# Tumour extrinsic neutrophil regulation in mesothelioma correlates with responsiveness to AXL and PD1 inhibition in MIST3, a phase IIA clinical trial

**DOI:** 10.64898/2026.09.22.26363540

**Authors:** Dean A. Fennell, Joanna Dzialo, Min Zhang, Aleksandra Bzura, Charlotte Poile, Essa Y. Baitei, Maurice Dungey, Amy Branson, Amy King, Shaun Barber, Marina Antoniou, Fatihme Maarawi, Maymun Jama, Liz Darlison, Molly Scotland, Clare Belcher, Amrita Bajaj, Bruno Morgan, Cassandra Brookes, Daniel Faulkner, Jan Rogel, Peter Wells-Jordan, Catherine Jane Richards, Jin-Li Luo, Emma Darlington, Louise Carter, Martin Little, Alastair Greystoke, Anne Thomas, Max Luckett, Mohammad Abdullah, Jens C. Hahne, Zisen Zhou, Hongji Yang, Sean Dulloo, Koirobi Haldar, Michael Barer, Gareth Griffiths, Sean Ewings, Kayleigh Hill, Eugene Tulchinsky, Nada Arabi Nusrat, Nathaniel C. Kuse, James Monkman, Rafael Tubelleza, Aaron Kilgallon, Arutha Kulasinghe, David Micklem, James B. Lorens, Matthew G. Krebs

**Author notes:** contributed equally Corresponding Authors details Dean A. Fennell FMedSci, FRCP, FRSB University of Leicester and University Hospitals of Leicester NHS Trust, Matthew G. Krebs, MD, PhD, Division of Cancer Sciences, Faculty of Biology, Medicine and Health, The University of Manchester C/O The Christie NHS Foundation Trust, Wilmslow Road, Manchester, M20 4BX, UK.

## Abstract

Immune checkpoint blockade (ICB) with ipilimumab and nivolumab is a front- line standard of care in patients with mesothelioma. However, only a minority of patients respond. The tyrosine receptor kinase AXL is a putative ICB resistance factor. We evaluated AXL and programmed death 1 (PD1) inhibition (AXL-PD1) with bemcentinib and pembrolizumab respectively, in a phase IIA clinical trial in patients with ICB naïve-relapsed mesothelioma (MIST3, NCT03654833). The study met its primary endpoint with a 12-week disease control rate of 46.2%, 90% confidence interval 29.2-63.8%. Grade 3 or greater adverse events occurred in 53.9% of patients. Multiomic analyses identified tumour-intrinsic and tumour extrinsic features linked to response to AXL-PD1 inhibition. Responders exhibited greater *NF2* loss, heightened immune response signalling, and epithelial mesenchymal transition, which was lower in biopsies obtained at the time of acquired drug resistance. Neutrophil- related transcriptional programmes were associated with response to AXL-PD1 inhibition. Neutrophil regulating erythroblast transformation specific (ETS) transcription factors SPI1, ETS2, FLI1 were differentially activated in responding tumours. Gut microbiota rheostat differed between responders and non-responders and positively correlated with neutrophil- associated transcription program, IRF3 and TLR2. In summary, our findings support a hypothesis linking gut microbiota composition, neutrophil- associated tumour biology and response to AXL-PD1 inhibition in mesothelioma.

## Background

Mesothelioma is a lethal cancer caused by exposure to asbestos that lacks effective therapy, particularly in the relapsed treatment setting ^1^. Recently, phase III clinical trials have confirmed the efficacy of immune checkpoint blockade (ICB) targeting programmed death 1 (PD1) ^2^ or in combination with either CTLA4 blockade ^3^, or chemotherapy ^4,5^. Despite these advances, the durability of immunotherapy remains relatively short. The mechanisms underpinning *de-novo* and acquired resistance to ICB remain under explored.

*De-novo* resistance to ICB has been linked to epithelial-mesenchymal transition (EMT) ^6^. EMT confers an immunosuppressive microenvironment and poor prognosis ^7–9^. AXL is an oncogenic receptor tyrosine kinase and member of the Tyro3 AXL Mer (TAM) family that is activated by growth arrest specific 6 (GAS6) ^10,11^. It is a key regulator of EMT – associated tumour plasticity ^12,13^.

Beyond its role in EMT, AXL has also emerged as immunomodulatory receptor with important functions in the tumour microenvironment^14–16^. Preclinical studies demonstrated that AXL inhibition can enhance responsiveness to PD-1 blockade through modulation of innate and adaptive immune responses^17^, while early clinical evaluation of bemcetinib combined with pembrolizumab in advanced non- small cell lung cancer demonstrated acceptable tolerability and preliminary antitumour activity^18^. Mesotheliomas express high levels of AXL ^19^. Inhibition of AXL reverses EMT ^20^ and promotes CD8^+^ T-cell-mediated antitumor immunity ^14,21,22^. Based on this, we explored the clinical efficacy of AXL inhibition by bemcentinib in combination with pembrolizumab (targeting PD1) in patients with immunotherapy-naïve, relapsed mesothelioma. Correlations between radiological response with both tumour intrinsic and extrinsic features were investigated, in order to determine potential mechanisms of *de-novo* and acquired resistance. Tumour extrinsic regulation of immunotherapy response by bacterial gut microbiota has been reported across several cancers including mesothelioma ^23–25^. Here we report the efficacy of combined AXL and PD1 inhibition (AXL-PD1) in patients with relapsed mesothelioma, revealing through multiomics, tumour intrinsic and extrinsic features that confer responsiveness.

## Results

### Patients

Between September 2020 and March 2022, 41 patients were assessed for eligibility to participate in MIST3, of whom 26 consented for treatment following screening (supplementary figure 1). The median follow-up time was 55.3 weeks (range, 4.9 to 150.3). Baseline patient characteristics are summarized in supplementary table 1. Median age of the cohort was 72.5 years (interquartile range (IQR) 69.0 – 75.0 years), of whom 23/26 (88.5%) were male, 23/26 (88.5%) had epithelioid histology, 8/26 (30.8%) had lymph node involvement, and 5/26 (19.2%) had metastases. Eastern Cooperative Oncology Group (ECOG) performance status was 1 in 20/26 (76.9%) patients. Most patients 20/26 (76.9%) reported exposure to asbestos. The majority 17/26 (65.4%) had previously received one course of systemic therapy; 7/26 (26.9%) had previously received two, and 2/26 (7.7%) had previously received three courses of systemic therapy (supplementary table 2). The complete list of eligibility criteria is summarized in the trial protocol in the supplementary materials.

All 26 patients who were clinically eligible for treatment received at least one cycle of treatment. The median number of cycles received of either drug within 24 weeks was 4.0 (range 1-8). The median time on study was 14 weeks, the reasons for discontinuation are shown in supplementary table 3.

### Efficacy

Disease control rate (DCR) at 12 weeks was assessed radiologically using modified RECIST at a scanning interval every 6 weeks, and was achieved in 12/26 patients (46.2%, 90% confidence interval (CI) 29.2-63.8%; 95% CI 26.6-66.6%). Partial response was observed in 4/26 (15.4%). Progressive disease rate was 11/26 (42.3%), and radiology was not evaluable in 3/26 (11.5%) due to early clinical progression and symptom burden precluding computed tomography (CT) re-evaluation. Radiological response was dichotomized into subgroups exhibiting no increase in tumour size (R, 17/24 shown in teal) *versus* tumour growth (NR, 7/24 shown in purple) (figure 1A). This response classification was based on best change in tumour size defined by mRECIST ≤ 0% versus >0%, respectively, and was subsequently used for R/NR comparisons in translational analyses. The translational cohort included one additional patient who was not evaluable for the primary endpoint but had available tumour measurements for best radiological response assessment.

**Fig. 1.**
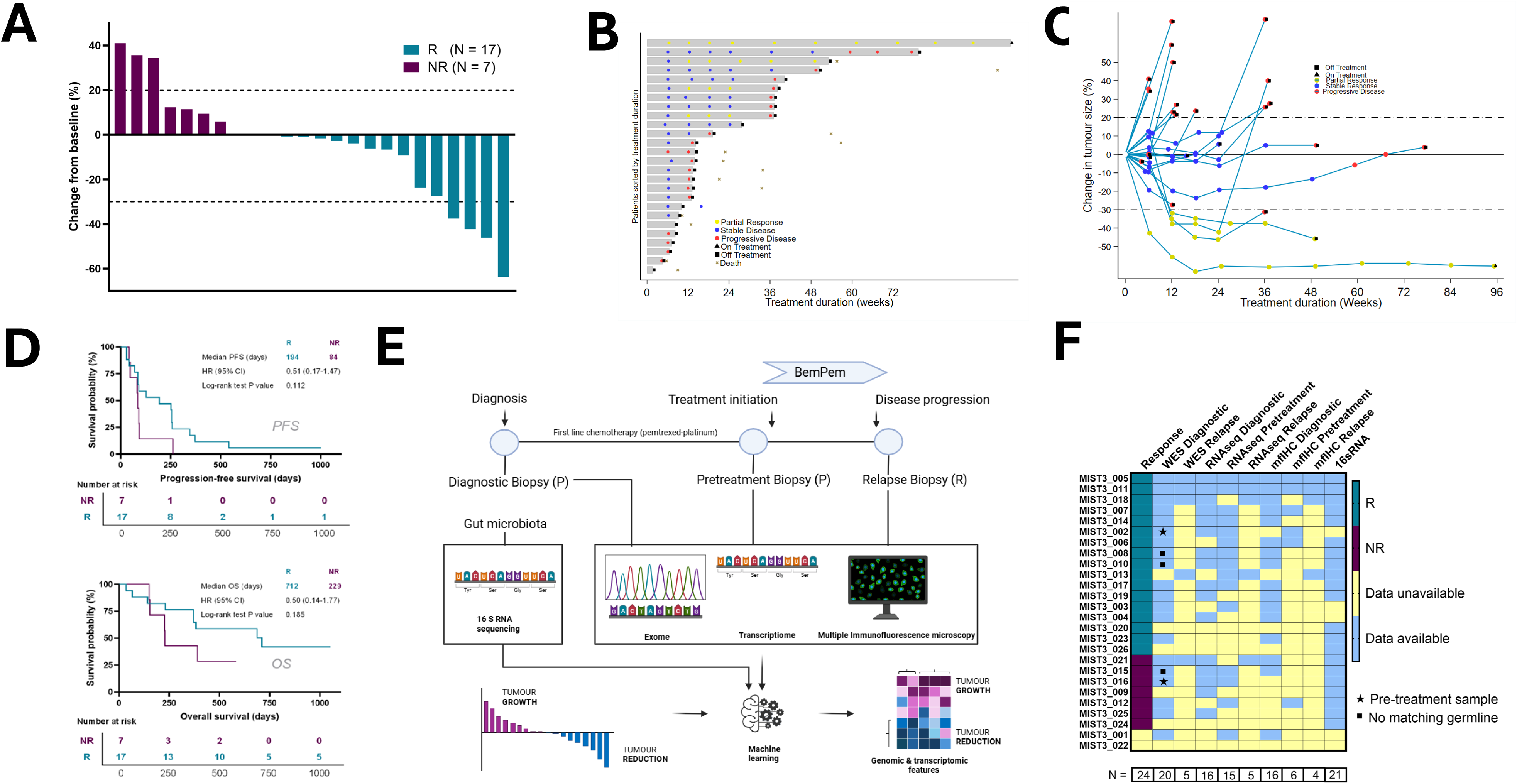
Efficacy of bemcentinib-pembrolizumab in patients with relapsed mesothelioma in MIST3 A. Waterfall plot showing best tumour response. Two patients progressed prior to baseline CT assessment. Dashed lines indicate modified RECIST1.1 thresholds for which the best change in tumour size (%) was either a partial response (PR) or progressive disease (PD). For the translational analyses, patients achieving disease control (*i.e*. % best change in tumour size equal or less than 0), are defined as R (cyan), those not meeting this criterion were called NR (purple). B. Swimmer plot of treatment course in MIST3. Each bar represents one patient; bar length corresponds to weeks on treatment. Periods on and off therapy and time of death are shown. Tumour response is shown, coded as yellow for PR, blue for stable disease (SD), and red for progressive disease (PD). The top patient remained alive and in treatment at data lock for the main analysis. C. Spider plot of tumour change (modified RECIST) over 24 weeks. Dashed lines denote thresholds for PR (-30%) and PD (+20%). D. Kaplan–Meier curves for PFS (top) and OS (bottom) stratified according to the post-hoc response categorisation (R versus NR) used in the translational analyses. Median durations in days, HR and log-rank p-values are shown. E. Overview of sampling and analyses. FFPE tissue collected at diagnosis (D), pre-treatment (Pre), and relapse after progression (Rb) underwent whole-exome sequencing, RNA sequencing and multiplex immunofluorescence. Gut microbiota profiling was performed using 16S rRNA gene sequencing. Machine learning approaches identified biological features enriched in tumours showing reduction or stability (R) *versus* growth (NR). F. Heatmap summarising translational cohort composition. Patients (in rows) are grouped by the translational response category (R vs NR) and ordered within each group according to sample availability. Columns represent individual data layers, and the number of samples available for each layer is indicated in boxes below the corresponding columns.

At 24 weeks 10/26 (38.5%, 95% CI 20.2-59.4%) had disease control (figure 1B-C). In a post*-hoc* analysis, patients were stratified according to translational response classification described above, where median progression-free survival (PFS) was 13.0 weeks (95% CI 10.0 – 36.3 weeks; and median overall survival (OS) was 55.3 weeks (95% CI 32.4 – 89.3 weeks). In the dichotomized groups R and NR, PFS and OS exhibited a trend to longer PFS (194 days *versus* 84 days, hazard ratio (HR) 0.5, log rank test p=0.185) and OS (712 days *versus* 229 days, HR 0.51, log rank test p=0.112) respectively (figure 1D).

### Safety and tolerability

At least one adverse event (AE) was experienced in 26/26 (100%) patients. Among these 26 patients, the most prevalent AE was fatigue in 12/26 (46.2%), followed by nausea in 11/26 (42.3%). Greater than one AE occurred in 24/26 (92.3%) of patients (supplementary table 4A). Of all AEs reported 171/248 (69.0%) were grade 1. Treatment-related AEs classed as possibly, probably, or definitely related to pembrolizumab occurred in 84/248 of AEs (33.9%) and related to bemcentinib in 101/248 of AEs (40.7%) (supplementary table 4B and 5).

Regarding AEs per individual, 12/26 (46.2%) had grade 1 or 2 as their highest grade of AE, 14/26 (53.9%) had grade 3 and 0/26 (0%) grade 4 or 5. Grade 3 AEs included colitis, diarrhoea, fatigue, urinary tract infection, hyponatremia, hypophosphatemia, arthralgia, pneumonitis, rash, joint pain, community acquired pneumonia, empyema, weight loss, thrombocytopenia, hypercalcemia, oral candidiasis, autoimmune hepatitis, and gallstones. AEs are summarized in supplementary table 6.

Serious AEs (SAEs) occurred in 10/26 (38.5%) patients, of whom 9/26 (34.6%) had one SAE, and 1/26 (3.9%) had two SAEs (supplementary table 7A). Dose delays occurred in 15/26 (57.7%) patients for pembrolizumab and in 23/26 (88.5%) patients for bemcentinib. Eight patients had a dose reduction to 100 mg bemcentinib during the trial period. SAEs led to permanent treatment discontinuation in 10/26 (38.5%), with 9/26 (34.6%) having SAEs that led to permanent treatment discontinuation of both pembrolizumab and bemcentinib and 1/26 (3.9%) that led to permanent discontinuation of pembrolizumab alone. SAEs deemed to be related to pembrolizumab occurred in 3/26 (11.5%), and for bemcentinib in 3/26 (11.5%, supplementary table 7B). A summary of all SAEs is shown in supplementary table 8.

### 22q loss is associated with sensitivity to AXL-PD1 inhibition

To elucidate tumour-intrinsic and extrinsic predictive factors associated with response in MIST3, a multiomic analysis was conducted as summarized in figure 1E. Whole-exome sequencing (WES) was performed on DNA extracted from diagnostic or pre-treatment mesothelioma formalin- fixed paraffin- embedded (FFPE) samples. Matched germline controls were available for 15 patients. WES sequencing depth averaged 200x for tumour, and 75x for germline DNA with 10-fold coverage >90%. In addition, WES was conducted on three post- treatment re-biopsy samples.

RNA sequencing was conducted in diagnostic (n=16), pretreatment (n=15), and re-biopsy (n=5) FFPE tissue blocks. Quality control data related to both DNA and RNA sequencing is provided in the supplementary data tables. The immune landscape was profiled using 49-plex immunofluorescence (n=16) and the gut microbiota profiled using 16S rRNA sequencing (n = 21, figure 1F).

Loss of chromosome 22q, a region containing multiple genes including the established mesothelioma driver *NF2,* encoding Merlin, was significantly enriched in the R- subgroup (Fisher exact test p-value 0.022, figure 2A-B, supplementary figure 2A). However, the genomic landscapes of R- and NR- mesotheliomas were similar with respect to cancer drivers, global mutation rate and burden, somatic copy number alterations, predicted neoantigens, uniparental disomy and homologous recombination deficiency mutational signatures (figure 2C).

**Fig. 2.**
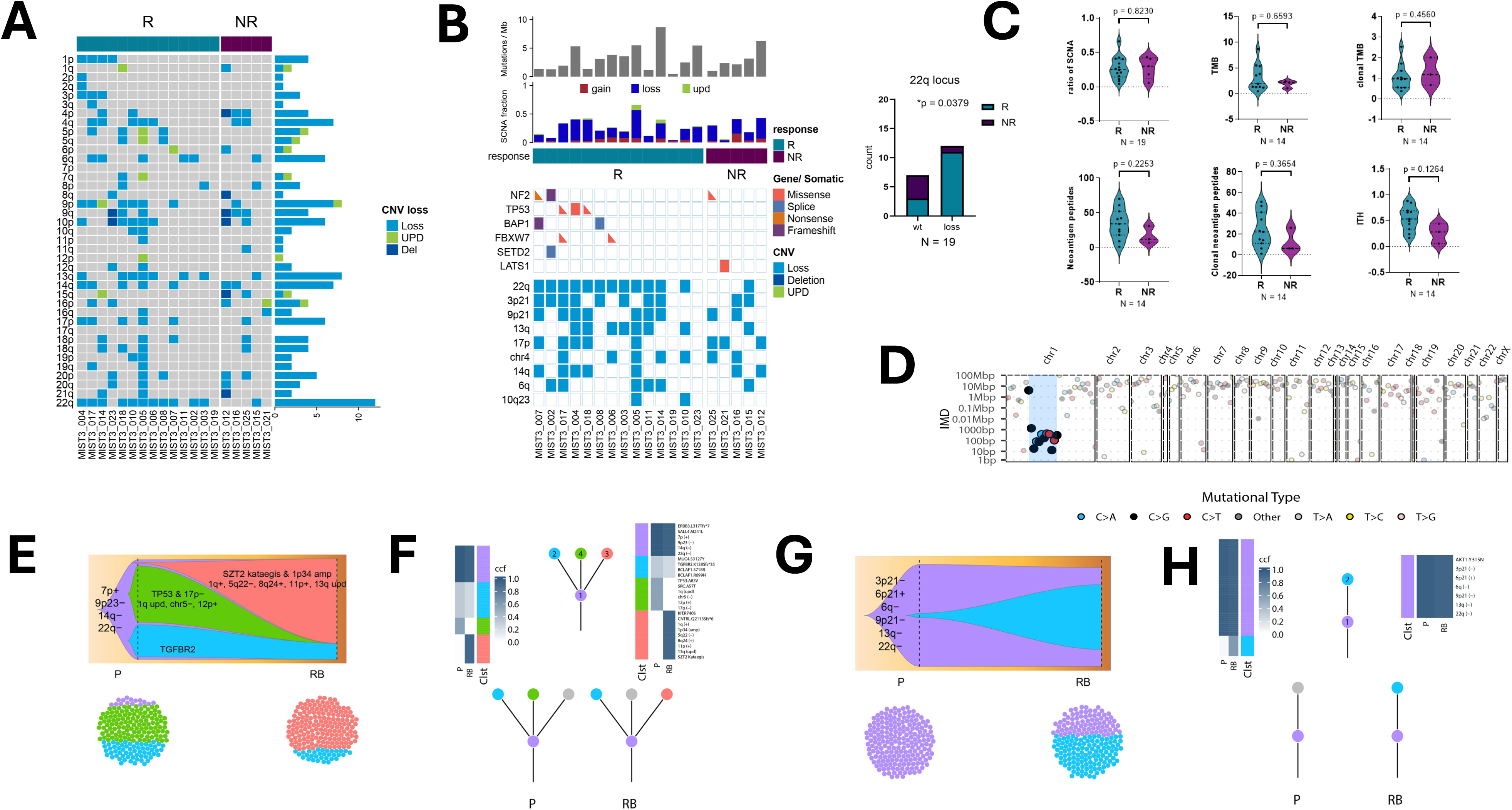
Genomic landscape and response in MIST3. A. Heat map showing the relative frequency of exome-wide chromosomal losses in the MIST3 cohort. B. Heatmap showing the relative frequency of single nucleotide variants and somatic copy number alterations involving cancer drivers in R *versus* NR patients. 22q loss was enriched in R (right panel; two-sided Fisher’s exact test, p value shown). C. Violin plots showing genomic features in responders (R, cyan) and non-responders (NR, purple). No significant differences were observed in SCNA ratio, total or clonal tumour mutational burden (TMB), neoantigen load, or intratumour heterogeneity (ITH) (two-sided Mann–Whitney test; p-values and sample sizes, n, are shown on the plots). D. Kataegis plot highlighting a localized hypermutation cluster on chromosome 1, coincident with 1q34 amplification, observed in the re-biopsy of MIST3-018 (absent at pre-treatment). E. Fishplot showing subclonal expansion of kataegis involving 1p34 amplification between pretreatment biopsy (P) and re-biopsy at resistance (RB) in patient MIST3-018. F. Clonal evolution model illustrating acquisition and expansion of a resistant subclone at re-biopsy (RB) in MIST3-018. G. Fishplot showing subclonal expansion of 9p21 loss at re-biopsy (RB) compared with pretreatment biopsy (P) in MIST3-011 H. Clonal evolution model depicting acquisition and expansion of a resistant subclone at re-biopsy (RB) in MIST3-011.

Longitudinal subclonal evolution was explored at the time of disease progression compared with the diagnostic timepoint. In patient MIST-018 the re-biopsy showed acquisition of subclonal kataegis involving 1p34 amplification consistent with an acquired catastrophic chromosomal event involving massive genomic instability (figure 2D-F, supplementary figures 2B-C). In patient MIST03_011, clonal dynamics analysis revealed expansion of a subclone harbouring losses at 9p21, 6q and 22q in the re-biopsy, despite a largely stable copy number profile between timepoints (figure 2G-H, supplementary figure 2D).

Gene translocations have been implicated as putative neoantigen driven immune responses ^26^, however the fusion frequency was not significantly different between the R- or NR-subgroups in either diagnostic or pretreatment mesothelioma biopsies (supplementary figures 3A-B).

### Neutrophil- associated transcriptional programme is enriched in responders to AXL-PD1 inhibition

R-mesotheliomas exhibited differential gene expression compared with NR- mesotheliomas. Gene set enrichment analysis (GSEA) employing the molecular signatures database (msigDB) hallmark collection ^27^ showed enriched inflammatory transcriptional signatures, notably interferons α and γ in the R-subgroup (figure 3A). EMT confers resistance to ICB in mesothelioma, as well as in pan-cancer analyses^28^, but this was not observed in MIST3 (figure 3A-B). To directly contrast this finding using the same analytical framework, an identical analysis was applied to combined data from two independent mesothelioma ICB cohorts (MIST4 and CONFIRM), revealing the opposite trend and recapitulating the previously reported association (figure 3C, supplementary figure 3C). At the time of disease progression, re-biopsy was conducted in five patients, followed by RNA sequencing to interrogate transcriptomic regulation at the time of drug resistance, and showed downregulation of EMT (figure 3D). To explore the immune landscape, transcriptome immune deconvolution was conducted using multiple established deconvolution approaches, generating immune-related scores and estimates for each sample, reflecting immune-cell abundance or activity. Correlation with response revealed myeloid lineage enrichment comprising neutrophils as the most highly associated cell type (Spearman’s r=- 0.87, permutation p=0.0002, figure 3E). CD8^+^ T-lymphocytes and monocyte/macrophages were also enriched in AXL-PD1 sensitive mesotheliomas shown by two or more deconvolution algorithms (figure 3E, supplementary figures 3D and 4). Given the involvement of CD8^+^ T cells in response to immune checkpoint inhibition, adaptive immune activity was further assessed using T- and B-cell receptor repertoire metrics. These features - including clonality, entropy, richness, and abundance inferred from RNA sequencing, were similar between R- and NR-groups in both diagnostic and pre-treated mesotheliomas (Wilcoxon p>0.05 supplementary figures 5A-F). As adaptive receptor usage showed no discriminatory value, subsequent analyses focused on the dominant myeloid signal.

**Fig. 3.**
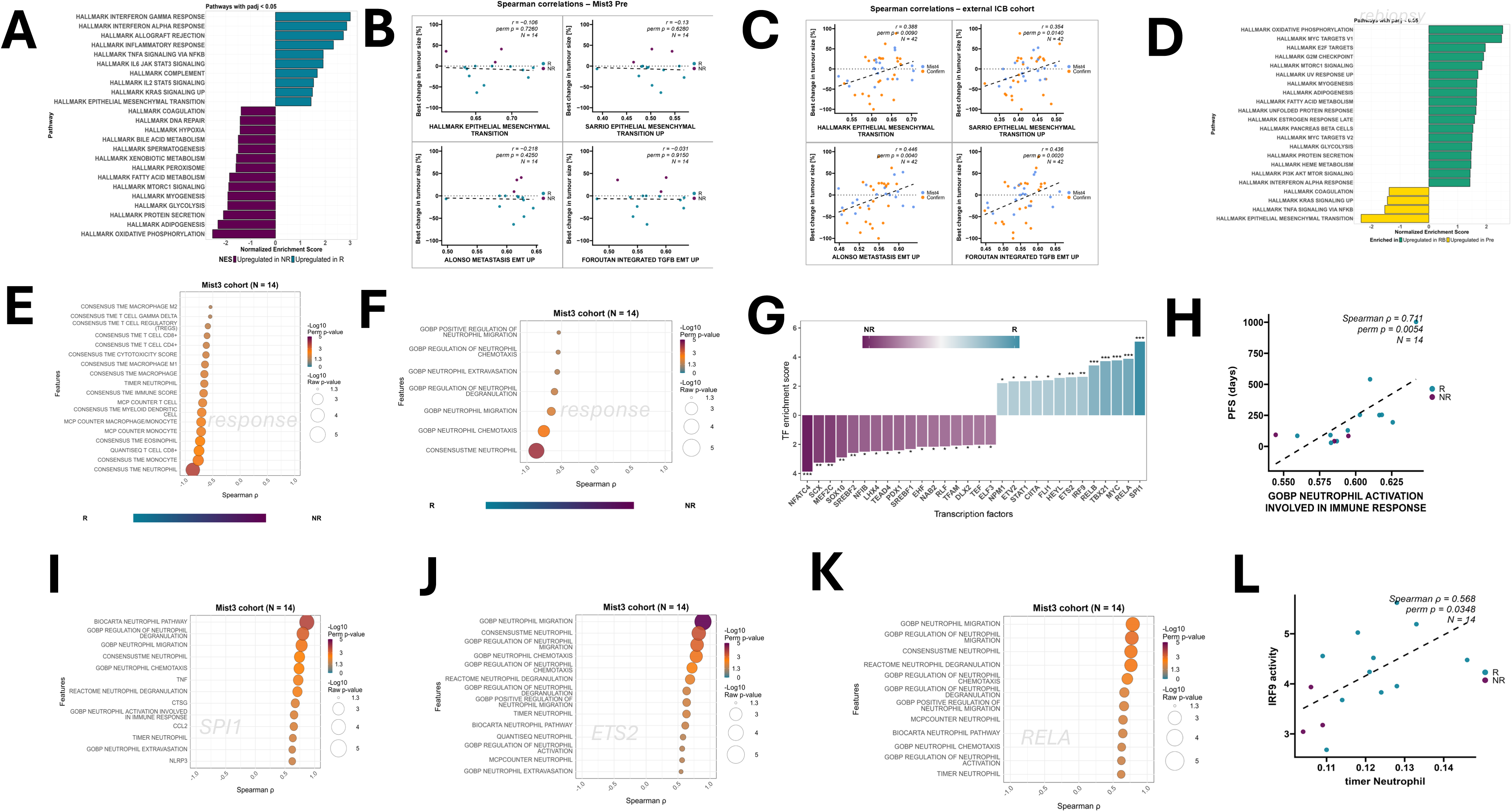
AXL inhibition may broaden ICI benefit in EMT-activated and neutrophil-inflamed mesotheliomas. A. Gene set enrichment analysis (GSEA) of pretreatment biopsies in MIST3 (N = 14). Bars show normalised enrichment scores (NES) for pathways differing between R and NR. B. Scatterplots showing the relationship between best tumour change and EMT-related ssGSEA scores in pretreatment MIST3 biopsies. C. Validation of EMT-tumour size relationships in the MIST4 and CONFIRM using the same EMT-related ssGSEA scores. Points are colour coded by cohort (MIST4 = blue; CONFIRM = orange). D. GSEA comparing pretreatment (Pre) and relapse (Rb) biopsies in MIST3 (N = 19). Bars show NES values. Positive NES (green) indicate pathways upregulated in Rb - relative to Pre, and negative NES (yellow) indicate pathways upregulated in Pre. E. Immune deconvolution of pretreatment biopsies in MIST3 using consensusTME, quanTIseq, MCP-counter, TIMER, and CIBERSORTx. Each bubble represents the correlation between one immune feature and best change in tumour size. F. Bubble plot showing correlations between neutrophil-related gene sets in MIST3 pretreatment biopsies and best change in tumour size. G. Transcription factor (TF) activities inferred in pretreatment biopsies (N = 14) using DecoupleR and CollecTRI – target interaction networks. Bars show NES values derived from model t-statistics. Significance: *p < 0.05, ** p< 0.01, *** p< 0.001. H. Scatterplot of ssGSEA scores for the gene set: GOBP Neutrophil activation involved in immune response versus progression-free survival (PFS) in pretreatment tissue from MIST3. I. Bubble plot showing correlations between SPI1 activity and neutrophil – related features in pretreatment biopsies. J. Bubble plot showing correlations between ETS2 activity and neutrophil – related features in pretreatment biopsies. K. Bubble plot showing correlations between RELA activity and neutrophil – related features in pretreatment biopsies. L. Scatterplot showing the correlation between IRF9 activity and TIMER- derived neutrophil estimates in pretreatment biopsies. R samples are shown in teal and NR in purple. Only significant correlations (raw or permutation p < 0.05) are displayed on bubble plots. Bubble size encodes -log10(raw p), and colour encodes - log10(permutation p). Sample sizes and statistical tests are indicated on plots or in the legend; permutation testing (10 000 label permutations) was used to confirm significance unless otherwise specified. For reference, -log10(p) = 1.3 corresponds to p ≈ 0.05 and is shown for orientation where applicable. The teal-purple bar in panels E-F aids interpretation of correlations with best change in tumour size by mRECIST (negative: R; positive: NR).

Further interrogation of neutrophil- associated transcriptomic signals showed strong correlations between tumour response and neutrophil chemotaxis, migration, and degranulation (figure 3F), while no such effect was observed in mesotheliomas treated with other immunotherapy regimens (supplementary figure 6A). Exploratory ROC analysis of the strongest response- associated neutrophil estimate supported the observed association with treatment response (bootstrap optimism-corrected AUC = 0.935; supplementary Fig. 6B). Consistent findings were also observed when response was analysed as continuous variable (C-index = 0.844, perm p = 0.0001; supplementary figure 6C).

Transcription factors SPI1, RELA, IRF9, ETS2 and CIITA associated with neutrophil- related programs were increased in the R-group, whereas NFATC4 showed the strongest relative decrease (figure 3G). A specific neutrophil activation- related gene program was associated with longer PFS in MIST3 (figure 3H). In exploratory analyses, both PFS and OS showed correlations with expression of neutrophil-linked chemokines and innate immunity signalling pathways, including CCL2, CXCL1 and toll-like receptor signalling (supplementary figure 6D). SPI1, which regulates myeloid, including neutrophil function was positively correlated with neutrophils associated transcription involving degranulation, chemotaxis, and activation (figure 3I). In addition, SPI1 was positively correlated with neutrophil regulatory factors CCL2, TNF, CTSG, and NLRP3 (figure 3I). Similarly, ETS2 and RELA showed broad correlations across neutrophil – related programs (figures 3J and 3K). IRF9 in turn showed a single dominant neutrophil – related association, best visualised with an individual scatterplot (figure 3L).

Interestingly, neither membranous nor cytoplasmic AXL protein expression were associated with response to AXL-PD1 inhibition (Spearman’s r=-0.289 permutation p=0.297 and Spearman’s r= 0.048 permutation p=0.866, respectively, supplementary figure 6E). Similarly, the plasma levels of AXL showed no association with response (Spearman’s r =0.138 permutation p = 0.521, supplementary figure 6F).

To complement transcriptomic findings, multiplex immunofluorescence was performed, to explore corresponding microenvironmental features at the protein level. This analysis identified stromal CD45RO, indicative of antigen experienced T-cell activation, as positively correlated with response to AXL-PD1 (Spearman’s r=-0.638 permutation p=0.012). In addition, vimentin, a marker of EMT (Spearman’s r=-0.609, permutation p=0.021), indoleamine 2,3-dioxygenase (IDO1) which promotes neutrophil infiltration (Spearman’s r=-0.559 and r=-0.617, permutation p=0.033 and 0.014 for tumour and stroma compartments respectively), and tumour proliferation rate (Ki67, Spearman’s r=-0.581 permutation p=0.023) were also positively correlated with tumour responsiveness (figure 4A-C). Conversely, the haemopoietic stem marker CD34 and CD141 (thrombomodulin) were significantly associated with resistance to AXL-PD1 inhibition (Spearman’s r=0.685, permutation p=0.0048 and Spearman’s r = 0.523, permutation p = 0.052 respectively, figure 4A-C). In common with previous reports^8,25^, CD8^+^ T-lymphocytes correlated with tumour responsiveness (Pearson’s r= -0.569, permutation p=0.0306, figure 4D). Ensemble machine learning comprising nine algorithms deconvoluted CD8^+^, CD4^+^ T-lymphocytes exhibited the highest average importance score, followed by granulocytes - the classical CD15/CD11b- associated phenotype (encompassing neutrophils), and CD14/CD68^+^ macrophage cluster consistent with tumour-associated macrophages expressing PD-1, HLA-DR and PD-L1 (figure 4E, supplementary figure 7A).

**Fig. 4.**
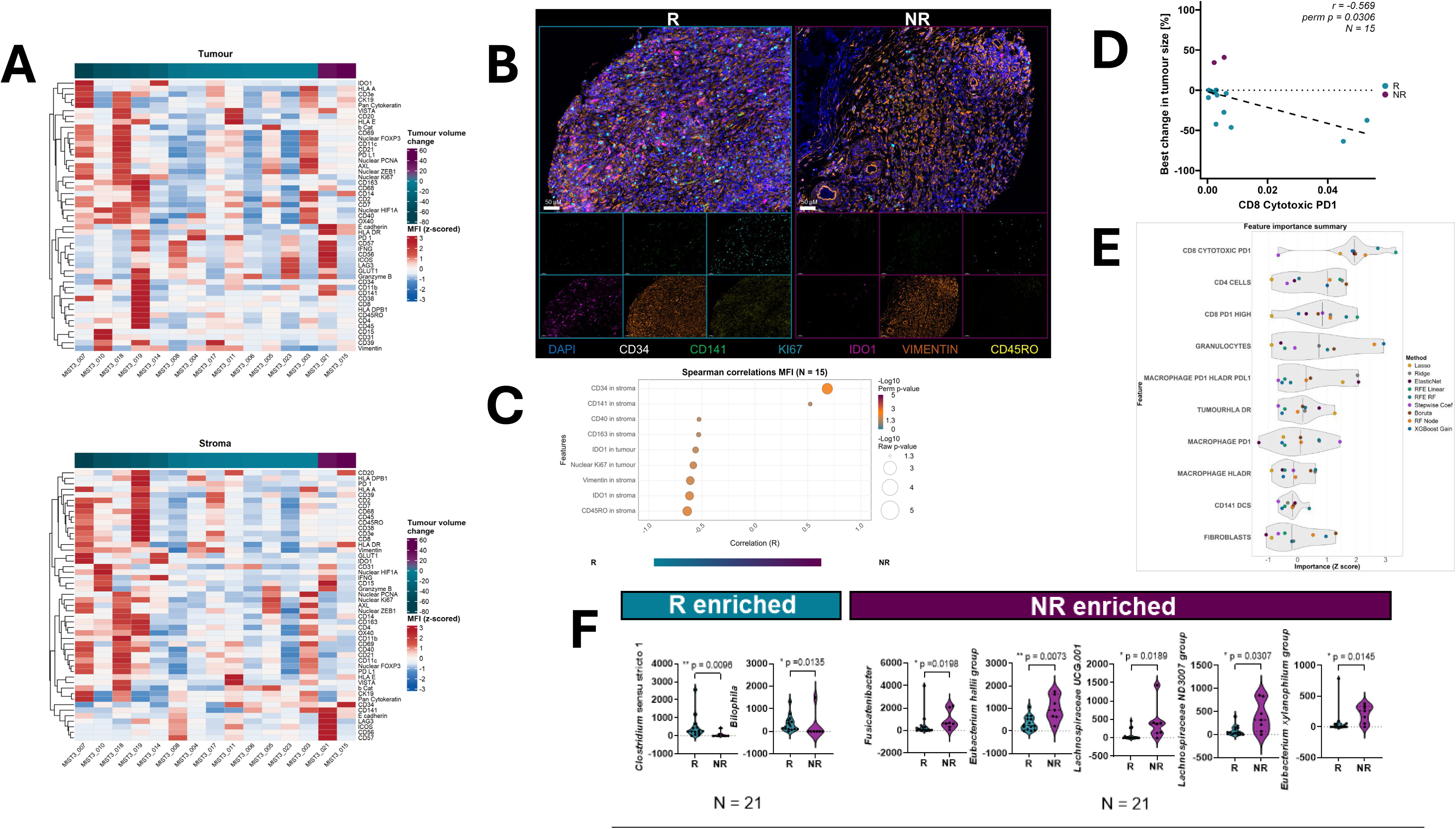
TME correlates of response and pretreatment microbiome differences in MIST3. A. Heatmaps depict z-scored mean fluorescence intensities (MFI) for 49 markers in the tumour (top) and stroma (bottom) compartments across diagnostic samples (N=15). Nuclear localization is shown for markers with nuclear signal; all others are quantified at the whole-cell level. Samples are ordered by best change in tumour size, and markers are hierarchically clustered by expression similarity. B. Representative multiplex immunofluorescence images for selected markers in responders (R, cyan) and non-responders (NR, purple). Composite images (top) include DAPI nuclear channel for orientation. Single-channel views (bottom) show each marker individually with identical scaling/contrast between R and NR. Markers were chosen based on their association with best percent change in tumour size (see Panel C). Images are exemplary. C. Spearman correlation analysis between marker expression (MFI) and percentage tumour- change across diagnostic samples (N=15). Each point represents one marker; only significant correlations (two-sided permutation and raw p < 0.05, 10 000 label permutations) are shown. Point size encodes −log10(raw p); colour encodes −log10(permutation p). The teal-purple bar aids interpretation of correlations with best percent change in tumour size (negative: R; positive: NR). D. Scatterplot showing the Pearson correlation between the CD8 cytotoxic PD-1 phenotype (from PRISM analysis) and best percent change in tumour size. Significance was assessed with 10 000 label permutations; the permutation p-value and Pearson’s r are annotated on the plot. Each point is one patient, coloured by clinical group (R = cyan, NR = purple). E. Multi-method feature selection of PRISM-derived phenotypes associated with best percent change in tumour size. Points show standardised importance (Z-score) from nine complementary selectors (Lasso, Ridge, ElasticNet, RFE-Linear, RFE-RF, Stepwise Coefficients, Boruta, RF node importance, XGBoost gain); violins summarize the distribution across methods with line at median. F. Pretreatment gut microbiome differences between R- and NR-mesothelioma patients (N=21) based on 16S rRNA gene sequencing. Genera were first nominated by Boruta on stratified R/NR groups, then confirmed by two-sided Mann–Whitney. Violin plots show relative abundances (points = patients; R = cyan, NR = purple, p-values from Mann-Whitney two-sided test displayed). Validated genera were then combined into Rheostat index (Log(G_R_/G_NR_)) for downstream integration with tissue features.

### Tumour extrinsic factors associated with neutrophil- linked response signatures

Gut microbiota composition has been reported to predict the sensitivity of ICB in a diverse range of cancers including mesothelioma ^25^. To determine if specific gut microbiota were associated with responsiveness to AXL-PD1 inhibition, 16S rRNA gene amplicon sequencing of gut microbiota was undertaken. Gut microbial diversity in the MIST3 cohort was similar in R- *versus* NR-subgroups as determined using eight methods Chao 1, dominance index, Goods coverage of counts, relative number of OTUs, Peilou’s evenness, Shannon index, Simpson’s index, and Faith’s phylogenetic diversity (supplementary figure 7B).

Using random forests machine learning, two bacterial genera, Clostridium sensu_stricto_1, and Bilophila were enriched in R- compared with NR-mesotheliomas. Conversely, Fusicatenibacter, Eubacterium hallii group, Lachnospiraacae UGG 001, Lachnospiracae ND3007 group and Eubacterium_xylanophilium group were all enriched in the NR- compared with the R-subgroup (Wilcoxon p<0.05 figure 4F, 5A, supplementary figure 7C). Using linear discriminant analysis (LDA) uncovered R- mesothelioma-associated gut microbiota enrichment of pathways linked to tryptophan biosynthesis, degradation of adenosine, guanine, glycerol, and L-1-2 propanediol degradation. Additionally, propanoate metabolism and glutathione peroxidase were also enriched in the R-subgroup (figure 5B).

**Fig. 5.**
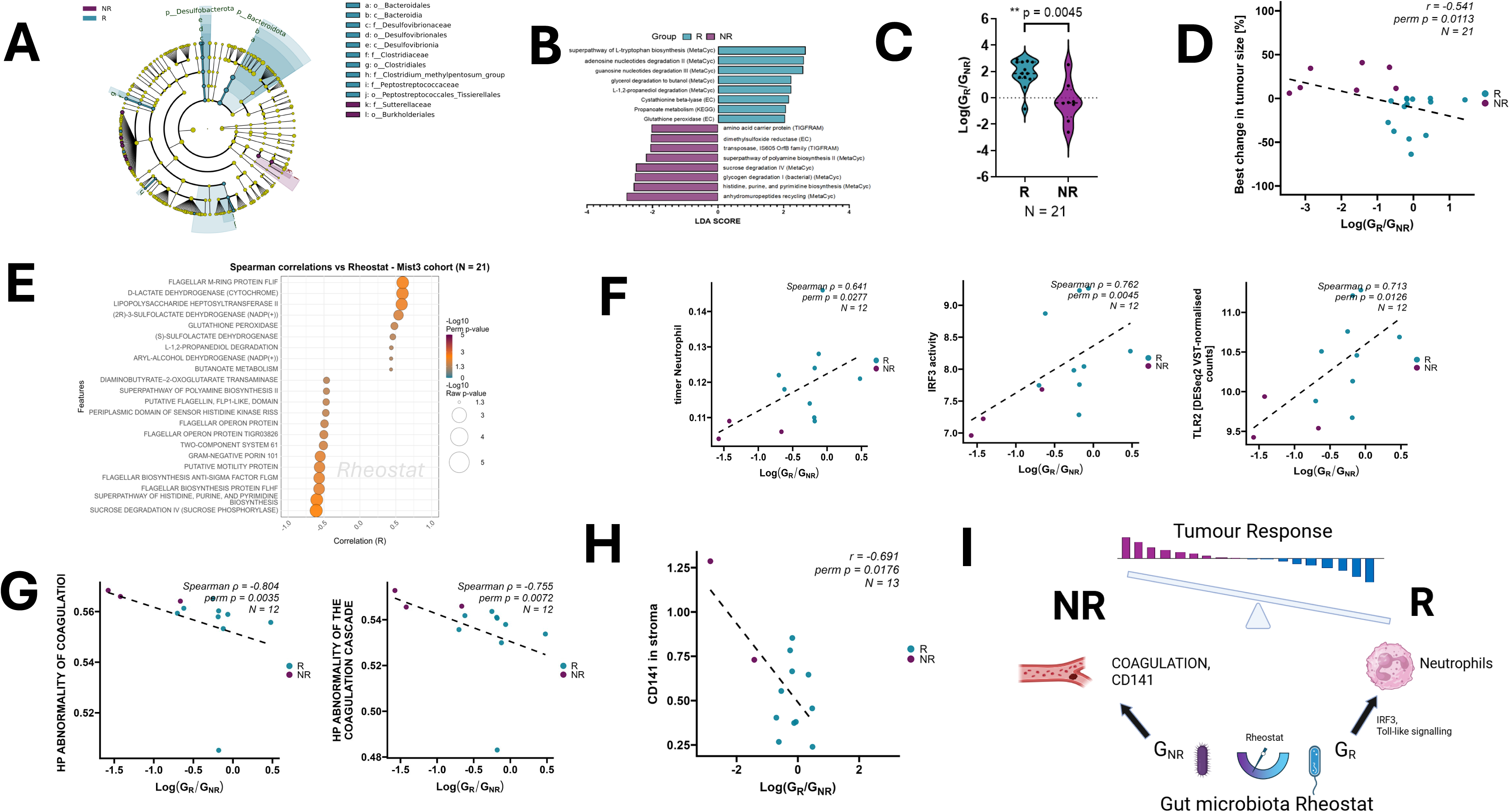
Gut microbiome –tumour axis associated with response to bemcentinib-pembrolizumab therapy. A. Circular cladogram showing phylogenetic relationships of the pretreatment gut microbiome in MIST3 (N=21). Colours indicate taxa enriched in R or NR. Concentric rings represent taxonomic ranks (phylum to genus). B. Predicted functional pathway differences inferred with PICRUSt2 and tested by LEfSe. Bars show LDA scores (log₁₀) for pathways enriched in R (cyan) or NR (purple). C. Violin plot of per-patient Rheostat index values in R (cyan) and NR (purple). Points denote patients. Group difference was assessed by two-sided Mann–Whitney test (p-value and N shown). D. Spearman correlation between Log(G_R_/G_NR_) and best percent change in tumour size. Each point is one patient. E. Correlations between the Rheostat and PICRUSt2- predicted functional pathways. Each bubble represents one pathway. Only significantly correlated pathways are displayed. Bubble size encodes -log_10_(raw p), and colour encodes -log_10_(permutation p). F. Scatterplots showing correlations between the Rheostat and selected tumour- intrinsic and immune -related features derived from pretreatment RNAseq. Each point represents one patient. G. Scatterplots showing correlations between the Rheostat and tumour- intrinsic, coagulation – related gene set signatures from MsigDB. Each point corresponds to one patient. H. Pearson correlation between stromal CD141 mean fluorescence intensity (MFI) in diagnostic tissue and the Rheostat. Points denote individual patients. I. Schematic summary of the gut-tumour axis suggested by the MIST3 analyses. Higher Rheostat values (R–enriched genera) associate with increased neutrophil infiltration and IRF3/Toll-like signalling features, whereas lower values (NR-enriched genera) associate with higher stromal CD141 expression and abnormal coagulation-related pathways observed in NR tumours. Together, these associations support a hypothesis-generating model linking gut microbial ecology, neutrophil-associated tumour features and response to AXL-PD1 therapy. R-samples are shown in teal, NR in purple across all panels; sample sizes (N) are displayed on the plots. Statistical tests (correlation and ROC analyses) are indicated in the legend or on the corresponding plots; permutation testing (10 000 label permutations) was used to confirm significance unless otherwise specified.

A MIST3 specific gut microbiota rheostat was computed as described previously ^25^ from R and NR- associated genera identified within the MIST3 cohort, based on the log ratio of the sum of amplicon sequence variant values corresponding to either R- or NR-mesotheliomas. The rheostat was associated with treatment response, demonstrating both differential distribution between R and NR groups (Mann- Whitney p= 0.0045; Figure 5C) and correlation with the best change in tumour size (Spearman r= -0.541, perm p= 0.0113; Figure 5D).

The gut microbiota rheostat was subsequently used as continuous representation of microbiome status for functional interrogation using PICRUST2- derived pathway analyses. Rheostat was positively associated with R-subgroup enriched pathways including glutathione peroxidase (Spearman’s r= 0.481, permutation p= 0.0271), L-1-2 propanediol degradation (Spearman’s r= 0.438, permutation p= 0.0487), and butanoate metabolism (Spearman’s r= 0.434, p= 0.0514). Flagellar M-ring protein FLIF was the most positively correlated bacterial feature (Spearman’s r= 0.596, permutation p=0.0046, figure 5E). Conversely, sucrose degradation was the most negatively correlated pathway with the rheostat (Spearman’s r= -0.609, permutation p= 0.0033, figure 5E).

The gut microbial rheostat was positively correlated with neutrophil infiltration, (Spearman’s r= 0.641, permutation p= 0.0277), interferon regulating factor 3 (IRF3) activity (Spearman’s r= 0.762, permutation p= 0.0045), TLR2 expression (Spearman’s r=0.713 permutation p=0.0126) (figure 5F). Conversely, abnormal coagulation signatures were the most negatively associated with the rheostat (figure 5G). Consistent with this finding, the rheostat was negatively correlated with CD141/thrombomodulin (Pearson’s r=-0.691, permutation p=0.0176; figure 5H).

## Discussion

Mesotheliomas rapidly acquire resistance to ICB, limiting therapeutic durability. AXL contributes to mesothelioma proliferation and invasiveness^19^, and confers an immunosuppressive tissue microenvironment (TME) ^13,29,14^. Targeting AXL reprogrammes the TME towards immune responsiveness ^16,21^, synergizing with PD1 blockade in preclinical models ^22,30^. On this basis, we explored the efficacy of AXL-PD1 in MIST3. MIST3 was a single arm phase II clinical trial, designed with the goal of identifying a potential signal of efficacy. A limitation of this study is the small sample size. The study met its primary endpoint, and validation would be required to confirm efficacy, through a follow on randomised phase III clinical trial. The translational analyses were also performed in a small cohort and should therefore be considered exploratory. However, integration of multiple data layers identified convergent tumour-intrinsic and tumour-extrinsic features associated with responsiveness to AXL-PD1 inhibition. External validation of these translational findings was not feasible within the current study, as no independent mesothelioma cohort treated with combined AXL- PD1 inhibition was available.

AXL induces EMT ^12,13^ which has been reported to correlate with resistance to immunotherapy in pan cancer and mesothelioma ^6,25,28^ cohorts. EMT is a pervasive phenotype in mesothelioma that exhibits a histo-molecular gradient ^31^. In the MIST3 cohort, EMT was not associated with resistance to AXL-PD1 therapy, in contrast to associations reported in other studies^8,25^. Indeed, loss of 22q/NF2 which enhances EMT *via* inactivation of the hippo pathway ^32–35^ was enriched in AXL-PD1 responsive mesotheliomas. Furthermore, the EMT-related protein vimentin was also overexpressed in R- *versus* NR-mesotheliomas ^36^. We observed downregulation of EMT in re-biopsies taken at the time of acquired resistance, and this may have been mediated by AXL inhibition, consistent with previous reports ^14,37^.

In MIST3, sensitive mesotheliomas harboured an inflamed tumour microenvironment, however clinical efficacy was not associated with a higher burden of predicted neoantigens, as was also previously reported in the CONFIRM (PD1 targeted) and MIST4 (VEGF-PD1 targeted) clinical trials ^8,25^. The recent identification of splicing aberrations involving GNAS resulting in a potent, public neoantigen ^38^ suggests that exome-level neoantigen prediction may fail to capture the full repertoire of TCR-bound peptides required to drive host anti-tumour immunity. In MIST3 responsive mesotheliomas exhibited enrichment of interferon α and γ, IRF9, and toll receptor-like transcriptional programs, reflecting a hot tumour microenvironment irrespective of the exomic total neoantigen burden.

Neutrophils were the most enriched immune cell type in AXL-PD1 responsive mesotheliomas in MIST3. Intratumour neutrophils exhibiting a SELL^hi^ state have been reported to accumulate in ICB responsive cancer ^39^. Tumour-infiltrating neutrophils, following chemotherapy have been reported to express GAS6, activating AXL and promoting tumour cell proliferation ^40^. Inhibition of AXL reverts the immunosuppressive phenotype through disruption of GAS6-AXL signalling ^22^. Our findings in MIST3 suggest that AXL inhibition may hijack protumour GAS6/AXL signalling emanating from neutrophils, reverting an immunosuppressive phenotype. Furthermore, tumour intrinsic upregulation of the transcription factor SPI1, required for terminal granulopoiesis ^41^, and ETS2, which regulates myeloid cell inflammation ^42^ were positively correlated with response to AXL-PD1 and tumour infiltrating neutrophils.

Gut microbial ecology influences responsiveness to ICB across multiple tumour types ^23,24,43,44^, including in mesothelioma ^25^. Short chain fatty acid (SCFA) metabolites produced by gut bacteria (butanoate, propionate) have been reported to alter tumour cell gene expression ^45^, and promote T-cell mediated antitumor immunity^46^. In MIST3, SCFA metabolism was associated with AXL-PD1 response-associated gut bacteria ^46^. Gut microbiota can also re-program tumour resident mononuclear phagocytes ^47^ and upregulate HLA class 1 ^48^. Beyond mononuclear phagocytes, gut-derived metabolites and bacterial vesicles can modulate other innate myeloid lineages; such signals have been reported to influence neutrophil recruitment and activation even in tumours located outside the gut ^49^. Here we show that the gut microbial ecology measured by a gut microbiota rheostat ^25^ was associated with both AXL-PD1 responsiveness and neutrophil infiltration. Interestingly, the microbial genera associated with response including enrichment of *Bilophila* and *Clostridium sensu stricto 1* in responders, and of the *Eubacterium hallii* group, *Fusicatenibacter*, and *Lachnospiraceae* subgroups in non-responders differed from microbial taxa previously reported to be associated with response to ICB in melanoma and NSCLC, including *Akkermansia muciniphila*^23^ and *Faecalibacterium*- associated communities^24^. The response- associated genera identified in the present study showed partial overlap with those reported in our previous mesothelioma cohort, MIST4, treated with PD-L1 and VEGF blockade^25^. These observations are consistent with reports across multiple cancer types demonstrating associations between baseline gut microbiota composition and immunotherapy outcomes, while highlighting the limited reproducibility of individual response-associated taxa across studies^43,50^.

In MIST3, tumour intrinsic coagulopathy was enriched in non-responders, and was inversely correlated with the gut microbiota rheostat. The tumour coagulome can regulate the tumour microenvironment ^51^; activated factor X has been reported to suppress antitumor immunity by signalling through protease activated receptor (PAR2)^52^. Thrombin mediates immune evasion *via* PAR1 ^53^. We observed a negative correlation between thrombomodulin (CD141) and the gut microbiota rheostat. CDC141 forms a complex with thrombin to activate protein C, which in turn downregulates proinflammatory genes and can directly bind to and inhibit neutrophil extracellular trap formation ^54^.

Our findings support a model derived from the observed associations, in which the gut microbiota may reciprocally modulate mesothelioma neutrophil infiltration and thereby contribute to responsiveness to AXL-PD1 inhibition (figure 5J). Previous studies have demonstrated that gut microbial ecology can be modified through dietary interventions ^50,55^ or microbiota transplantation ^56^, may promote a favourable gut bacterial ecology to facilitate responsiveness to immunotherapy. However, as these observations are derived from correlative multiomics analyses, the proposed microbiota-neutrophil axis should be viewed as a hypothesis-generating model that requires further validation in future clinical and translational studies.

In summary, mesothelioma responsiveness to AXL-PD1 inhibition was associated with a gut microbiota-neutrophil axis and inversely associated with coagulome- related features, but not EMT, highlighting the potential importance of tumour-extrinsic TME regulation as a targetable enhancer of immunotherapy efficacy in this rare cancer setting.

## METHODS

### Study design and participants

We conducted MIST3, a multi-centre, one-stage, single arm open label phase IIa study at three UK centres; the University Hospitals of Leicester National Health Service Trust, and the Northern Centre for Cancer Care, Newcastle upon Tyne, and The Christie NHS Foundation Trust, Manchester.

Patients were eligible for the study if they were aged 18 years or older and had histologically confirmed mesothelioma that had shown radiological progression after at least one completed course of standard first line systemic treatment with pemetrexed and either cisplatin or carboplatin. Patients with pleural mesothelioma could be enrolled irrespective of the histological subtype. Any line of treatment was permitted (excluding any prior immunotherapy) with prior therapy completing no less than 14 days before treatment was initiated.

Patients were required to have measurable disease by modified Response Evaluation Criteria in Solid Tumours for malignant mesothelioma (mRECIST1.1), predicted life expectancy of 12 weeks or more, ECOG performance status score of 0–1, adequate haematological (full blood count including total white cell count, neutrophils, platelets and haemoglobin), renal (urea and electrolytes), and liver function tests (including bilirubin, alkaline phosphatase, alanine transaminase) and willingness to undertake research blood tests and optional tissue re-biopsy for translational research (please refer to the protocol in the Supplementary Information).

Exclusion criteria included diagnosis or treatment of any other cancer within the 5 years before study entry, treatment with any agent with no marketing authorization within 30 days before study entry, and palliative radiotherapy in the 4 weeks before baseline CT scan, uncontrolled brain metastases, and cardiac, respiratory, hepatic or renal insufficiency (full exclusion criteria are given in the appendix pages 36-40). The protocol (included in the Supplementary Information) was approved by the East Midlands Leicester South Research Ethics Committee (reference 18/EM/0118), and the Medicines and Healthcare products Regulatory Agency (MHRA). The study was registered on ClinicalTrials.gov (NCT03654833).

During MIST3 pre-screening, archival diagnostic FFPE tissue blocks collected as part of routine clinical care were accessed for molecular analyses within MiST platform. In addition, patients consented to undergo protocol-defined pretreatment CT- guided tumour core biopsies prior to initiation of treatment. In those patients who exhibited disease control exceeding 12 weeks, an optional second tumour core biopsy at disease progression was obtained on a subset of eligible patients. Additional tissue analysis was conducted under research ethics approval 14/LO/1527, a translational research platform entitled Predicting Drug and Radiation Sensitivity in Thoracic Cancers. The study was further approved by University Hospitals of Leicester NHS Trust (reference IRAS131283) with the University of Leicester being a sponsor.

The study was completed in accordance with the Declaration of Helsinki and the principles of Good Clinical Practice as defined by the International Council for Harmonisation (IHC). Written informed consent was obtained from all patients before enrolment.

### Procedures

Patients were eligible if they had progressed following at least one course of prior systemic treatment for mesothelioma that included standard first-line pemetrexed and either cisplatin or carboplatin. Oral bemcentinib was given once daily in 21-day cycles, initially loaded with three days of 400 mg, then from day 4 onwards 200 mg once daily was administered; pembrolizumab was given intravenously at a fixed dose of 200 mg on day 1 of a 21-day cycle, together for a period of 24 weeks. This dosing schedule was based on prior clinical experience with bemcentinib and was consistent with previous clinical studies evaluating bemcentinib in combination with pembrolizumab^18^. Response was assessed by CT scan every 6 weeks until week 24; thereafter, CT scans were done every 12 weeks. Radiological response was evaluated according to mRECIST1.1^57^ by investigator-assessed review. Tumour response assessments were based on a local review *(i.e.,* no central review of response data). Patients who had disease control beyond 24 weeks could continue to receive bemcentinib and pembrolizumab outside the trial through the UK’s Medicine and Healthcare Products Regulatory Agency (MHRA) individual patient access mechanism until disease progression, unacceptable toxicity, or withdrawal of consent. Any patient not receiving a single dose of the study drug was excluded from the efficacy population and replaced until all 26 efficacy evaluable patients were recruited (please refer to the full protocol in the supplementary materials).

Patients had safety monitoring visits (including physical examination, blood tests, and toxicity assessments) at the start of each cycle, with additional visits on day 15 of cycles one and two, and follow-up visits 30 days and 6 months after the last dose (Supplementary Information). Dose interruption was allowed for National Cancer Institute Common Terminology Criteria for Adverse Events (NCI CTCAE; version 4.03) grade 3 or 4 toxicity, for a maximum of 12 weeks, until complete recovery or reversion to grade 2 toxicities. A single dose reduction to bemcentinib was allowed: corresponding to 100 mg (dose level –1), once a day. Dose escalations were not permitted. Patients who discontinue pembrolizumab (for reasons other than disease progression) were able to continue with bemcentinib monotherapy, no dose modifications to pembrolizumab were permitted.

### Outcomes

The primary endpoint was disease control rate (DCR) at 12 weeks, defined as the number of patients with complete response, partial response, or stable disease, as a proportion of the total number of patients who received at least one dose of the study drug. The 12-week landmark was used to establish a threshold for activity and has been frequently used as a primary endpoint in phase 2 studies in mesothelioma ^25^. There is currently no standard of care in the relapsed disease setting. In the placebo group of the negative Vantage phase 3 trial (comprising 332 patients) ^58^ median PFS was 6 weeks. Consequently, we estimated that a 12-week disease control rate of 50% would approximate to a doubling of expected PFS (for placebo), indicating a potentially useful treatment. Secondary endpoints were the safety and toxicity profile, DCR at 24 weeks, and best objective response rate. Objective response rate was defined as the proportion of patients whose best overall response was complete or partial. Safety was assessed by the incidence of AE, reported according to the NCI CTCAE, version 4.03.

### MIST3 statistical analysis

We used a single-stage A’Hern design with a type 1 error rate (one-sided) of 0.05 and power of 80%. The 12-week DCR parameters were set at *p*_o_ = 0·25 (*i.e.*, a true disease control rate of 25% at 12 weeks would be too low, requiring no further evaluation therefore accepting the null hypothesis) and *p*_1_ = 0·50 (*i.e*., a true disease control rate of 50% at 12 weeks would be sufficient to warrant further evaluation). These parameters required a total of 26 evaluable patients to be analysed. On the basis of these assumptions, if 11 or more of the 26 enrolled patients achieved disease control at 12 weeks, we would conclude that the criteria for success had been met. The efficacy population was defined as all patients who received at least one dose of the study drugs. The primary outcome was analysed in the efficacy population; we calculated the DCR at 12 weeks with exact two-sided 90% confidence intervals (CIs). All secondary endpoints and safety outcomes were analysed in the efficacy population. The disease control rate at 24 weeks and objective response rate with exact (two-sided) 95% CIs were calculated. Serious adverse events and adverse events were summarized by number, event, frequency, outcome, treatment given, severity (grade), and investigator-assessed relatedness to bemcentinib and pembrolizumab.

Safety and toxicity outcomes were determined for the safety population, defined as all participants who received at least one dose of trial medication. Primary and secondary analyses were done after all patients completed 24 weeks of treatment, or at withdrawal. Safety reports of patients who were on treatment beyond 24 weeks, up to 6 months, are provided to the drug provider separately. Categorical variables were summarized by frequencies and continuous variables were summarized by medians with interquartile ranges (IQRs).

In a post-hoc analysis (not protocol specified), progression-free survival was measured in weeks from the first dose to the date of progressive disease or death from any cause, censoring patients at the last known study visit assessment without evidence of disease or death. OS was measured in weeks from the first dose to the date of death from any cause, censoring all other patients at data lock. Median PFS and OS were estimated using the Kaplan–Meier method, and for comparison of survival curves Mantel–Cox *p*-value < 0.05 was considered significant. MIST3 statistical analyses were performed using STATA version 16.0.

### Formalin-fixed paraffin embedded (FFPE) tissue assessment and processing

FFPE tissue biopsy blocks were used for nucleic acid (DNA and RNA) extraction. After sectioning haematoxylin and eosin (H&E) stained slides were examined by a histopathology advanced biomedical scientist with support from a consultant histopathologist, who identified and marked representative areas of tumour on the H&E slides. From the diagnostic FFPE tissue blocks multiple tissue cores (1.0 mm each in size) were taken from the marked areas and DNA/RNA sequentially isolated using the MagMAX^TM^ FFPE DNA/RNA Ultra Kit (ThermoFisher Scientific, Waltham, MA, USA #A31881) on the Kingfisher^TM^ Flex sample purification system (ThermoFisher Scientific, Waltham, MA, USA) as per manufacturer’s instructions.

DNA was quantified using the Qubit™ 1× dsDNA HS assay (ThermoFisher Scientific, Waltham, MA, USA #Q33230) and RNA was quantified using the Qubit™ RNA HS assay kit (ThermoFisher Scientific, Waltham, MA, USA Q32852) on the Qubit™ 4.0 fluorometer (ThermoFisher Scientific, Waltham, MA, USA) according to manufacturer’s instructions. Whereas for the pretreatment and relapse FFPE tissue blocks, the material was obtained from tumour-marked areas identified on tissue sections. DNA/RNA extraction was then performed using the Qiagen AllPrep DNA/RNA FFPE Kit (Qiagen, Hilden, Germany, #80234). Quantification and quality assessment of extracted RNA were performed by the sequencing service provider using standard spectrophotometric and electrophoretic methods. DNA quality assessment was carried out using the Qubit™ 1× dsDNA HS assay (ThermoFisher Scientific, Waltham, MA, USA #Q33230).

### Germline DNA extraction

Germline DNA was isolated from buffy coat samples using the QIAamp DNA Blood Mini Kit (Qiagen, Hilden, Germany #51104). DNA was quantified using the Qubit™ 1 x dsDNA HS assay (ThermoFisher Scientific, Waltham, MA, USA #Q33230) on the Qubit™ 4.0 fluorometer.

### Whole exome sequencing

All diagnostic, pretreatment and relapse biopsy samples were processed through the same DNA sequencing provider and analytical pipeline. A small number of low-input samples were processed using an alternative method to accommodate limited material. A total amount of 1.0 μg genomic DNA per sample was used as input material for the DNA sample preparation. Sequencing libraries were generated using the Agilent SureSelect Human All ExonV6 kit (Agilent Technologies, San Diego, CA, USA) following the manufacturer’s recommendations and index codes were added to each sample. Briefly, fragmentation was carried out by a hydrodynamic shearing system (Covaris, Woburn, MA, USA) to generate 180–280 bp fragments. Remaining overhangs were converted into blunt ends via exonuclease/polymerase activities and enzymes were removed. After adenylation of 3′ ends of DNA fragments, adaptor oligonucleotides were ligated. DNA fragments with ligated adaptor molecules on both ends were selectively enriched in a PCR reaction. After PCR reaction, library hybridize with Liquid phase with biotin labelled probe, after which streptomycin-coated magnetic beads are used to capture the exons of genes. Captured libraries were enriched in a PCR reaction to add index tags to prepare for hybridization. Products were purified using AMPure XP system (Beckman Coulter, Beverly, CA, USA) and quantified using the Agilent high-sensitivity DNA assay on the Agilent Bioanalyzer 2100 system. Qualified exome capture libraries were then sequenced on the Illumina NovaSeq 6000 platform (Illumina, San Diego, CA, USA), according to standard protocols, for 150 bp paired-end multiplexed sequence.

### Processing of WES sequencing data

After removing sequencing reads with low quality and adaptor bases using FASTP, clean reads were aligned to human reference genome (UCSC hg19) using Burrows-Wheeler Aligner (bwa-0.7.17). Mapped genomes were sorted using Sambamba (v0.6.7). Duplicate reads were marked using Picard tools (v2.18.9). Somatic SNVs and INDELs were detected with VarScan2 and MuTect2[2] jointly. Briefly, VarScan2 somatic (v2.3) were used to do somatic variants calling with default parameters, except for the following: minimum coverage for normal and tumour sample were set to 10 and 8 separately, minimum variant frequency was adjusted to 0.01 and tumour purity was set to 0.5. As to MuTect2 dealing process, we used MuTect2 contained in GATK bundle (4.0.5.1), with default parameter. ANNOVAR was used for functional annotation of variants.

### SNV and INDEL filtering

To reduce false positive variant calls, further filtering strategies were used on the mutation detection results of both MuTect2 and VarScan2. A variant was retained when it was both detected by MuTect2 and VarScan2 (somatic *p*-value ≤ 0.1 for SNV and ≤0.05 for INDEL) with vaf >2% or only detected by VarScan2 with a VAF > 5%. In matched tumour sequencing data, filtering thresholds were defined based on variant allele frequency (VAF) and read support (VAF threshold: 1%, alternative read count thresholds: 5 for SNVs and 2 for INDELs). Besides, variants located on the regions of simple repeats and segmental duplications were also removed. The population frequency of SNV did not exceed 1% in any of the following population-based database − 1000 Genome, EXAC or ESP6500. An additional filter was applied to exclude artefact mutations introduced by the preparation of FFPE specimens and sequencing libraries, which are characterized as the bias of the variant read support by DKFZBiasFilter.

### SCNA calling, tumour purity and ploidy estimation

We use ASCAT to estimate somatic copy number alterations (SCNA) in paired tumour tissue/normal tissue sequencing datasets. The estimated tumour purities and ploidies were also corrected by manual review of ABSOLUTE results. Allele counts of positions from 1000 genomes were generated using AlleleCounter, and minimum coverage of 20 for normal sample was used for filtration. LogR and BAF values were produced for each region and concatenated into one matrix separately for each patient. LogR values were subsequently corrected using a GC wave correction implemented in ASCAT, and only heterozygous BAF values were reserved for further analysis. Allele-specific segmentation was performed to generate segmented logR and BAF data by ascat.aspcf. Manual verification was used to select the optimal model for ploidy and cellularity using an orthogonal measure based on ABSOLUTE results and mutation variant allele fraction. And then ASCAT was re-run to obtain the final allele-specific copy number data using reviewed cellularity and ploidy.

### HRD signature analysis

Homologous recombination deficiency (HRD) scores are determined using the scarHRD R package. HRD score based on allele-specific copy numbers is sum of loss off heterozygosity (LOH), telomeric allelic imbalance (TAI), large-scale transitions (LST) scores. HRD-LOH score is the number of 15 Mb exceeding LOH regions which do not cover the whole chromosome. HRD-TAI is allelic imbalances that extend to the telomeric end of a chromosome. HRD-LST is defined as chromosomal break between adjacent regions of at least 10 Mb, with a distance between them not larger than 3 Mb.

### Clonality analysis

A modified version of PyClone was used to estimate the cancer cell fraction (CCF) of the mutations and perform clustering analysis. For a given mutation we first calculated the observed mutation copy number, nmut, describing the fraction of tumour cells carrying a given mutation multiplied by the number of chromosomal copies at that locus using the following formula (1):

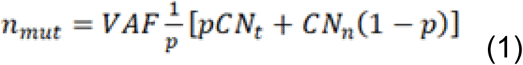

where VAF corresponds to the variant allele frequency at the mutated base, and *p*, CNt, CNn are respectively the tumour purity, the tumour locus specific copy number, and the normal locus specific copy number (CNn was assumed to be 2 for autosomal chromosomes). We then calculated the expected mutation copy number, nchr, using the VAF and assigning a mutation to one of the possible local copy number states using maximum likelihood. In this case only the integer copy numbers were considered. Mutations were then clustered using the PyClone Dirichlet process clustering. For each mutation, the observed variant count was used, and reference count was set such that the VAF was equal to half the pre-clustering CCF. Given that copy number and purity had already been corrected, we set the major allele copy numbers to 2 and minor allele copy numbers to 0 and purity to 0.5; allowing clustering to simply group clonal and subclonal mutations based on their pre-clustering CCF estimates. We ran PyClone with 10,000 iterations and a burn-in of 1000, and default parameters, with the exception of --var_prior set to ‘BB’ and –ref_prior set to ‘normal’.

### HLA typing and HLALOH

HLA typing for MHC class-I genes was carried out using POLYSOLVER (v1.0) software for all 28 normal samples’ bam files, with default parameters. In brief, reads in the WES data potentially originate from HLA gene region were extracted out and then aligned to genomic sequence library of all known HLA alleles based on IMGT, using Novoalign packaged in POLYSOLVER. After which, a two-step Bayesian classification approach was used to infer the two alleles for each HLA class-I genes (HLA-A, HLA-B and HLA-C). A crucial part of neoantigen presentation is the HLA class-I gene products, which can present tumour associated epitopes to T-cells and then trigger an adaptive immune response. Loss of heterozygosity in HLA genes may lead to decreased ability to present productive tumour neoantigens, which could facilitate immune evasion of cancer. LOHHLA software was used to evaluate HLA loss for all tumour samples, based on the alignment results of both tumour and corresponding normal samples, inferred tumour purity and ploidy information, and the HLA class-I genotyping results detected above. In brief, HLA reads were extracted and re-aligned to the patient-specific HLA-I alleles, then HLA gene specific log ratio was calculated based on coverage information on mismatch positions between homologous HLA alleles, and finally, HLA haplotype specific copy number was determined. In the analysis, items with PVal_unique ≤0.01 (difference in log ratio between allele 1 and allele 2 ≤ 0.01) were considered as a LOH event.

### Neoantigen prediction

Neoantigens were defined as 8–11-mer peptides resulted from somatic SNVs or InDels which led to amino-acid changes and, binding affinity score between remodelled peptide and respective patient’s HLA class-I molecules was <500 nM. Somatic mutation VCF files both from VarScan2 and Mutect2 were annotated by Variant Effect Predictor (Version 84) with default parameter, except for the using of ‘downstream’ and ‘wild type’ plugins offered by pVACseq53. After annotation, the variants leading to peptide changes were extracted out for downstream analysis. Bam-readcount (0.8.0) was used to acquire sequencing-based read depth information on each selected variant for both tumour and matched normal samples. Annotated non-synonymous mutations, sequencing-based information as well as HLA class-I gene typing results inferred by POLYSOLVER were feed into pVACseq(4.0.9) for neoantigen pre-diction. For each pVACseq run, epitope prediction was done by both NetMHC and NetMHCpan algorithms packed in pVACseq toolkit, epitope length was set to 8–11 and tumour DNA VAF cut-off was set to 10, with default parameters used for all other settings. Epitope prediction was performed based on the selected prediction algorithms, after which, sequencing-based information was integrated to enable filtering of neoantigen candidates (Normal Coverage ≥5×, Normal VAF ≤ 2%, Tumour Coverage ≥10×, Tumour VAF ≥ 10%). Inferred neoantigen candidates were selected out and those with binding affinity fold change >2 were considered with higher priority level, which means the ratio of binding affinity score between wild-type peptide and mutated peptide. The greater this value, the stronger of the binding affinity after mutation compared with wild-type epitope.

### RNA sequencing

RNA sequencing for diagnostic and pretreatment/relapse samples was performed by different service providers using distinct RNA purification and library preparation workflows.

For diagnostic samples, total RNA underwent ribosomal RNA depletion using a standard probe-based removal workflow. The rRNA-depleted RNA was then used for strand-specific cDNA library construction using a NEBNext® Ultra^TM^ RNA Library Prep Kit for Illumina® (NEB, Ipswich, MA, USA). After adaptor ligation, amplification and size selection to enrich for fragments of approximately 150-200bp, libraries were quality assessed and pooled for sequencing using paired-end reads.

For pretreatment and relapse tissue derived RNA, samples were prepared using RNA exome capture-based library construction workflow. Following RNA fragmentation, sequencing adaptors were ligated and libraries were amplified prior to hybridization with capture probes targeting exonic regions of the transcriptome. Captured libraries underwent purification and quality assessment before pooling for sequencing, which was performed using paired-end reads (2x76bp) to a target depth of approximately 50 million read pairs per sample.

For both workflows, Illumina platforms were used for sequencing. FASTQ files were processed using the same computational pipeline for alignment, quantification and downstream analyses. Fastp (0.12.2) was used to remove low-quality reads and reads containing sequencing adapters. RSeQC (v5.0.3) was employed to perform quality assessment of RNA sequencing. The processed reads were aligned using STAR (2.6.1d) onto the human genome reference (UCSC hg19), and the transcripts were annotated based on gencode V19 gene models. Only the reads unique to one gene and which corresponded exactly to one gene structure were assigned to the corresponding genes by using HTSeq. STAR-Fusion (1.9.0)^59^ was applied to predict gene fusion events from RNA-seq data.

### Immune repertoire analysis

We applied TRUST4 (v1.0.0) to obtain TCR and BCR clonotypes from bulk-RNA-seq data for each sample. Raw pair-end reads were aligned to hg19 TCR/BCR sequences, and candidate reads were extracted to perform de novo assembly on V, J, and C genes including the hypervariable complementarity determining region 3 (CDR3). The assembled consensus sequences were re-aligned to IMGT reference gene sequences for annotation. The statistics of TCR and BCR, including abundance, richness, Shannon entropy and clonality, were compared between reduction and no-reduction groups using the Kruskal–Wallis rank-sum test.

### Immune deconvolution

RNA-seq gene expression data, normalized as transcripts per million (TPM), were analysed using several immune deconvolution algorithms to estimate immune and stromal cell composition. Five methods suitable for tumour microenvironment analysis were implemented from the immunedeconv R package^60^ with default settings: QuanTIseq, MCP-counter, TIMER, EPIC and ConsensusTME. In addition, CibersortX^61^ was run using the LM22 reference signature in absolute mode with 500 permutations.

Outputs from all algorithms were combined into a single data matrix, containing estimated immune and stromal cell fractions or scores for each sample and each method, and were used for downstream analyses.

### Gene set enrichment analysis

Gene set enrichment analysis was conducted on MIST3 pretreatment samples, MIST3 relapse samples and validation samples. Validation cohort consisted of data obtained from two previously published immunotherapy trials (MIST4^25^ and CONFIRM^8^). Gene expression count tables were used as input data and processed the in same way as MIST3 samples, with inclusion of batch correction for analyses involving validation data.

The Gene Set Enrichment Analysis (GSEA) was performed using the fGSEA R package^62^. Differential expression statistics were generated with DESeq2, and genes were ranked using Wald test statistics prior to enrichment testing. Multiple testing correction was applied using the Benjamini–Hochberg false discovery rate (FDR) method and adjusted p-value <0.05 was considered significant. For analyses including validation cohorts, cohort was included as batch covariate in the DESeq2 design.

In addition, the GSVA package^63^ was used to perform single-sample GSEA (ssGSEA) on DESeq2 variance stabilized (VST) normalized counts to estimate pathway activity per sample. VST normalization was used to ensure consistency across datasets and enable analyses across multiple cohorts.

ssGSEA included the Hallmark collection from MSigDB^27^, together with project-relevant gene sets compiled from several MSigDB categories, including immune response, tumour microenvironment, metabolic and cell- cycle related pathways. Complete list of gene sets from custom gmt file is included in supplementary information. The resulting enrichment scores were used for downstream marker analysis as well as for longitudinal analysis of pathway dynamics.

### Transcription factor activity analysis

TFs activity was inferred using the decoupleR package in R (v2.9.1)^64^. The Univariate Linear Model (ulm) was fitted for each TF and sample, where the observed gene expression serves as the response variable and the TF-target interaction weights act as explanatory variables. Information on TF- target interactions and regulatory effects (activation or inhibition) was obtained from the CollecTRI database. To compare aggregate differences in TF activity between responder (R) and non-responder (NR) groups, the ULM model was also run using moderated t-statistics from the differential expression analysis as input. The resulting enrichment scores represent the model - t- values, reflecting the relative activation or repression of each TF between groups. P-values were used to assess the significance of associations and TFs with p < 0.05 were considered significant.

### AXL immunohistochemistry and plasma AXL levels

AXL-IHC staining of FFPE-tumour sections was performed on an automated Ventana tissue stainer (anti-AXL clone 7E10, Thermo Fisher Scientific, Waltham, MA, USA) and scored according to study-specific guidelines by a trained pathologist at Roche Tissue Diagnostics (Tucson, Arizona, USA) as previously reported ^65^. Tumour-cell AXL expression (tAXL) was quantified via “Histo-score” (Hscore), which ranges from 0-300 and represents the sum of the percentages of cancer-cell staining at each intensity level multiplied by the cytoplasmic staining intensity (0=no staining, 1=weak- staining, 2=moderate-staining, and 3=strong-staining). Sections were scored for the abundance of total and AXL-expressing tumour-infiltrating immune cells (Total IC and AXL IC, respectively) as a percentage of all cells within the tumour area Pretreatment plasma concentrations of soluble AXL were measured using a validated multiplex immunoassay (Custom HumanMAP panel) performed by an accredited external service provider according to their standard operating procedures. Results were reported in ng/mL.

### Tissue Microarray (TMA) design and preparation

Patients enrolled in the MIST3 trial provided diagnostic formalin-fixed, paraffin- embedded (FFPE) blocks (D=diagnosis). Additionally, many patients had pre- treatment biopsies (P=pre-treatment) and biopsies at disease progression (R=relapse). TMAs were prepared externally according to the service provider’s standard procedures. A competent individual assessed the quality of H&E stained sections, marking suitable areas for the TMA. Maps were designed to ensure even distribution of triple cores per donor block. Where available, tissue from more than one biopsy type was included in the TMA block. The TMA block was manually produced using a Beecher Instruments device. To maintain structural integrity during sectioning, the precise positions for punching cores in the recipient block were determined using calibrated X and Y micrometers. Tissue punches from donor blocks were accurately placed into corresponding holes in the recipient block according to the TMA map.

### Multiplex staining

Staining was conducted by the Akoya Biosciences (Marlborough, MA, USA) research team following their optimized protocol. The list of markers used is presented in the supplementary data tables. TMA sections were submerged in two changes of xylene, followed by an ethyl alcohol gradient and water. Heat-induced antigen retrieval was performed, and the slide was incubated with water, hydration buffer, and staining buffer. A cocktail of antibodies conjugated with barcodes, diluted in staining buffer with a mix of four blocking solutions, was applied. After cover slipping with parafilm, the slide was incubated overnight at 4^O^C. Post-incubation, the slide was washed with PBS and fixation solutions (PFA and methanol) and stored in storage buffer until ready for imaging. The plate with barcodes was prepared for the PhenoCycler (Akoya Biosciences, Marlborough, MA, USA) run, placing one-three reporters recognizing appropriate antibodies in the wells of the reporter plate corresponding to the predefined imaging cycles. A flow cell was attached to the slide and placed into the flow cell slide carrier, which was run in the PhenoCycler-Fusion system.

Reporters are applied to the tissue by the instrument and visualized through fluorescence microscopy using the PhenoImager component of the PhenoCycler-Fusion Instrument. Each run consists of multiple cycles: in each cycle, reporters reveal up to three markers (plus DAPI), the tissue is imaged in different fluorescence channels, and reporters are removed with an isothermal wash. Repeating these cycles with various reporters allows visualization of the complete PhenoCycler antibody panel in a single experiment on the same tissue area.

### Image Analysis

TMA images were analysed using QuPath v0.5.1^66^, an open-source software with a broad range of tools for multi-step image analysis. Pre-processing steps included automated TMA de-arraying, tissue detection, annotating regions of interest, and cell detection with sparse (1 µm) area around nuclei reserved for cytoplasm. Object classifier based on artificial neural network algorithm (ANN_MLP) was trained to recognize tumour and stroma areas. Per cell measurements were exported from QuPath and processed as follows: average values were calculated for each tissue annotations (TMA core), then median was taken for repeats of the same patient, the same timepoint samples and data were obtained as mean fluorescence intensity per patient for both tumour and stroma segments.

In parallel phenotyping analysis was conducted. TMA scan was imported into QuPath where TMA cores were dearrayed. Partial, folded or missing cores were excluded from analysis. Cell segmentation was performed using Cellpose v3 plugin^67^ using nuclear model on DAPI channel with 4um cell expansion constrained at 1.7 times nuclear size. Mean expression per channel and nuclear morphology measurements were exported to csv and parsed into Anndata in python 3.10. Subsequent clustering by phonograph^68^, cell type annotation, neighbourhood generation, and cell frequency were performed within PRISM python package^69^. Expression matrices were arcsinh transformed, followed by initial round of clustering on immune and epithelial markers to separate tumour cells from non-tumour cells. Non-tumour cells were then clustered using cell lineage markers including CD45, CD3e, CD8, CD4, FOXP3, CD20, CD68, CD14, CD31, CD34, CD15, CD141, CD11B. Clustering was performed in parallel for leiden resolutions 0.1 to 0.5 with a range number of neighbours (K) 15-60. Clustering quality was evaluated by the adjusted rand index (ARI) and the davies-bouldin index. The final clustering parameter (K= 15, r=0.3) resolved 14 clusters which were annotated according to canonical lineage marker expression (supplementary figure 5A). Cell types included B-cells, CD4^+^ T-cells, CD8^+^ T-cells, CD4^+^ Tregs, CD141^+^ dendritic cells, granulocytes, macrophages, fibroblasts, blood vessels, tumour cells and a population of unidentified cells. Further functional classification was performed on CD4^+^, CD8^+^ and macrophage cells for PD1, PDL1, granzyme B and HLA-DR. Marker positivity was refined into distinct functional cell populations including CD4^+^ PD1^+^ TFH cells, CD8^+^ PD1^+^ exhausted cells, CD8^+^ granzyme B cytotoxic cells, as well as Macrophage populations positive for PDL1, HLA-DR, and PD1. Tumour cells were functionalised by AXL and ZEB1 positive marker expression. Cell frequencies were calculated by the proportion of each cell type within the whole tissue, or tissue region, relative to their respective total number of cells.

For downstream correlative analyses, diagnostic tumour samples were used, as these were the most consistently available across the cohort.

### Feature Selection Analysis

Feature selection was performed to identify tissue phenotypes and cell populations derived from PRISM analysis that were most strongly associated with clinical response.

To assess the importance of each cell phenotype for response prediction, a multi-method machine learning pipeline was employed, implementing diverse model families to minimise algorithm-specific bias. The following feature selection methods and R packages were applied:

- Lasso and Ridge, ElasticNet using glmnet package^70^: ElasticNet was run with alpha systematically varied from 0.05 to 0.95 in increments of 0.05, to avoid convergence to pure lasso (alpha=1) or pure ridge (alpha=0); for Lasso and Ridge, default settings were used.
- Recursive Feature Elimination (RFE): performed using the caret package^71^, with the base estimator set as either a linear model (RFE-linear) or a random forest (RFE-RF; using caret`’`s internal interface to the randomForest package); default caret settings were used, except where manual ranking adjustments was made for comparability with other methods; a scores of zero were assigned to all non-selected features prior to z-score normalization.
- Stepwise Model Selection (Stepwise Coef): the MASS package was applied with default settings, unless stepwise feature selection failed, in which case the feature importances were extracted only from successful runs; non-selected features were assigned a value of zero, ensuring all features were represented in the cross- method harmonisation.
- Boruta was run using Boruta package^72^ with maxRuns = 2000 and missing value imputation enabled, to further increase robustness and reduce random effects, the procedure was repeated 10 times (with different random seeds) and final feature importance scores were calculated as the mean importance across all runs; features rejected as unimportant were assigned a value of zero prior to aggregation and normalization.
- Random Forest and Node Importance (RF Node) was implemented using randomForest package^73^ with default settings.
- XGBoost Gain applied xgboost package^74^ with default parameters for feature importance computation.

For each method, feature importance scores were extracted, with z-score normalization applied for comparability. The distribution of standardized importance for each feature across all selection algorithms was summarized and visualized using violin plots, with individual points reflecting method-specific scores and line representing median score per feature.

### 16S rRNA gene sequencing and gut microbial community analysis

Stool samples for 16S rRNA sequencing were collected prior to initiation of treatment using ISO13485:2016 accredited sample collection kits (Atlas Biomed, London, UK). 16S rRNA genes in 16S V3-V4 regions were amplified with specific and barcodes. The 16S primer sequences are proprietary to Atlas Biomed and therefore confidential. All PCR mixtures contained 15 μL of Phusion® High-Fidelity PCR Master Mix (New England Biolabs, Ipswich, MA, USA), 0.2 μM of each primer and 10 ng target DNA., After PCR composed of 30 cycles at 98 °C (10 s), 50 °C (30 s) and 72 °C (30 s) and a final 5 min extension at 72 °C the PCR products were analysed on 2% agarose gel. Finally, PCR products were purified with Qiagen Gel Extraction Kit (Qiagen, Hilden, Germany) following manufacturer’s recommendations. NEBNext® UltraTM IIDNA Library Prep Kit (New England Biolabs, Ipswich, MA, US Cat No. E7645) was used for generating sequencing libraries and library quality was evaluated on the Qubit@ 2.0 Fluorometer (ThermoFisher Scientific, Waltham, MA, USA) and Agilent Bioanalyzer 2100 system (Agilent Technologies, Santa Clara, CA, USA). Finally, the library was sequenced on an Illumina platform (NanoSeq Illumina, San Diego, CA, USA) and 100 bp single-end reads were generated.

### Gut microbial community analysis

Fastp (-q 19 -u 15 -n 25 -l 60 -min_trim_length 10) software was used to do quality control of single-end raw data and to generate high-quality sequencing reads. Vsearch software was used to blast clean reads to the silva (v138) database to detect the chimera and remove them, so as to obtain the final effective data, namely effective tags. For the effective tags, the deblur module in QIIME2 software was used to do denoise, and the sequences with an abundance less than 5 were filtered out to obtain the final ASVs (Amplicon Sequence Variables) and feature tables. Then, the Classify-sklearn module in QIIME2 software was used to compare ASVs with the database and to obtain the taxonomical classification of each ASV. The read counts of ASVs were normalized to the sequence counts of the sample with the lowest sequencing depth. The taxonomic abundances of samples were summarized according to the ASV species annotations. Specificity was 97% to the genus level and only 80% to species level. The Boruta algorithm was employed to discern genera that exhibit significant associations with either the R or NR subgroups. A *p*-value < 0.05 was carried out using the Mann–Whitney test, illustrated using box plots of relative microbial abundance. A microbiota rheostat was computed from the logarithm (base 10) of the ratio of the sum of the relative abundances corresponding to significant genera in the R subgroup (*G*_R_) divided by the sum computed for NR-subgroup (*G*_NR_), i.e.

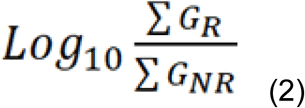

Hereafter, the rheostat is referred to as log(*G*_R_/*G*_NR_) in the manuscript. Spearman’s rank correlation coefficient was used to determine the association between log(*G*_R_/*G*_NR_) and tumour-intrinsic features, taking a *p*-value of <0.05 as significant.

### Microbiota diversity

Gut microbial alpha diversity was computed in QIIME2 using 8 methods; Among them, 1. chao1 (Chao1 index), 2. dominance (Berger-Parker Dominance index), 3. observed_otus (Number of distinct features) a richness index, 4. goods_ coverage (Good’s coverage of counts), 5. pielou_evenness (Pielou’s evenness) a measure of coverage and relative evenness of species. 6. shannon (Shannon’s index) and 7. simpson (Simpson’s index) an indicator of microbiota diversity, and 8. faith_pd (Faith’s phylogenetic diversity), also anther diversity index that incorporates phylogenetic difference between species.

### Gut microbiota - Linear discriminant analysis effect size

To identify significantly different bacteria between the two responses to immunotherapy at the genus level, LEfSe (version 1.0.8)^75^ was performed using the default setting. Significance was set at *p*-value < 0.05 and LDA score cut-off point of 2. A cladogram representative of the structure of the R and NR microbiota was generated.

Additionally functional metagenomic predictions were generated from ASV tables using PICRUSt2^76^, producing gene family and pathway abundance profiles across KEGG enzyme (EC), KEGG pathways, TIGRFAM, PFAM and MetaCyc databases. LEfSe analysis was applied to these functional profiles to identify metabolic pathways discriminating R and NR groups, with LDA scores quantifying the effect size and indicating the direction of enrichment.

### Machine learning

Random forest classification was used to select relevant variables. The importance of each variable was compared with the maximum importance value of all random (shadow) features using a permutation test. Analysis was repeated 10 times using 5000 iterations each. The R statistical software version 4.1 and the’ranger’ package were used for random forest training and variable importance elimination.

### Integrated translational analyses

To mitigate the impact of modest sample size and imbalance between R and NR groups, continuous measures were used where feasible. Dichotomised R/NR comparisons were applied for data types not readily amendable to continuous modelling.

Continuous variables were compared between response categories using the Mann – Whitney U test (two – sided, unpaired, significance threshold p < 0.05). Associations between categorical variables were evaluated with the Fisher’s exact test (two – tailed). Correlations between continuous variables were assessed using either Spearman’s rank or Pearson’s correlation coefficients. To support robustness of association testing in the modest-sized cohort, empirical p values were additionally obtained by permutation testing in R (10000 permutations, significance threshold for both tests p < 0.05).

Receiver operating characteristic (ROC) analyses were performed in R using the pROC package^77^. For each binary comparison (positive class = R), the empirical ROC curve with automatic orientation, the area under the curve (AUC), and 95% confidence intervals were computed. Statistical significance of discrimination was evaluated with a two-sided permutation test against AUC = 0.5 (10000 permutations; p < 0.05). To assess robustness of ROC- derived associations, bootstrap optimism correction and continuous-response concordance index (C-index) analyses were performed where feasible. ROC results were independently verified in GraphPad Prism for validation. Survival analyses were performed in GraphPad Prism using Kaplan–Meier estimation, and differences between the groups were assessed by the logrank estimation of median time to event parameters (overall and progression-free survival). *P*-values were estimated using Mantel–Cox testing with a *p*-value of <0.05 as the threshold for significance. Cox-proportional regression analysis was used to calculate hazard ratios. Post-hoc Analyses employed R version 4.3.2 and Prism 9.5.1 (Graphpad, San Diego, CA, USA). Illustrations were created with Biorender.com.

In view of the high-dimensional nature of the datasets, findings supported by multiple analytical approaches were prioritised during interpretation.

## Data Availability

Due to patient confidentiality and ethical restrictions, individual participant data are not publicly available but may be available from the corresponding author upon reasonable request and subject to appropriate approvals.

## Acknowledgements

The research was carried out at the National Institute for Health and Care Research (NIHR) Leicester Biomedical Research Centre (NIHR203327). For the purpose of open access, the author has applied a Creative Commons Attribution license (CC BY) to any Author Accepted Manuscript version arising from this submission. The MIST study is sponsored by the University of Leicester (Leicester, UK) and funded by the Asthma + Lung UK and British Lung Foundation Partnership and the Victor Dahdaleh Foundation (Toronto, ON, Canada; grant VPDCF17-17). The study drugs (bemcentinib and pembrolizumab) were provided by BerGenBio (Bergen, Norway) and MSD (New Jersey, USA) respectively. Additional funding is provided by Cancer Research UK in conjunction with the UK Department of Health on Experimental Cancer Medicine Centre grant (C10604/A25151). We thank Adrian Nicholson, Judith Underwood, Azmina Sodha-Ramdeen (University Hospitals of Leicester NHS Trust, Leicester, UK), Dr Caroline Cowley (Cancer Research Centre, University of Leicester, Leicester, UK), and Sam Moody (The Northern Cancer Research Centre, Newcastle, UK) for their valued contribution to this study. Furthermore, we acknowledge the Advanced Imaging Facility at the University of Leicester (RRID:SCR_020267) for their support. We especially thank the patients and their families who were involved in this clinical trial, and Mesothelioma UK. Matthew Krebs acknowledges support from National Institute for Health Research (NIHR) Manchester Biomedical Research Centre (NIHR203308), NIHR Manchester Clinical Research Facility at The Christie (NIHR203956) and Manchester Experimental Cancer Medicine Centre (Manchester, UK). Jake Spicer was funded by an MRC Doctoral Training Partnership iCASE studentship. Maurice Dungey is an NIHR clinical lecturer. Mohammad Abdullah and Max Luckett were supported by Wellcome Trust Biomedical scholarships.

## Reporting summary

Further information on research design is available in the Nature Portfolio Reporting Summary linked to this article.

## Data availability

Patient data related to this clinical trial shall remain confidential to the sponsor organisation (The University of Leicester) and will not be disclosed except where disclosure might be required in accordance with pharmacovigilance duties of the parties involved. Individual participant data can be made available, after deidentification to investigators who provide written request in accordance with General Data Protection Regulation and following authorization from the sponsor organization, starting immediately and ending 3 years after publication. Requests for data and materials will be reviewed by the sponsor and any implications regarding intellectual property or confidentiality considered. Raw sequencing data have been deposited in the European Genome-phenome Archive (EGA) under study accession EGAS50000001704 and are available through controlled access in accordance with patient consent and data protection requirements. All of the other data supporting the findings of this study are available within the article and its supplementary information files and from the corresponding author upon reasonable request. Source data are provided with this paper.

## Author contributions

D.A.F. and M.G.K. conceived the study.

D.A.F., M.G.K, A.Branson, A.King., S.B, P.W.J., A.T., C.J.R., L.D., E.D., L.C., M.Little, A.G. contributed to the clinical study design

D.A.F., M.G.K., C.P., J.D, E.Y.B, P.W.J., C.J.R., D.M., J.B.L., A.Branson, A.King, contributed to the translational study design

D.A.F., M.G.K., A.Bzura, M.Z., E.Y.B., J.D., C.P., M.D., J.R., P.W.J., C.J.R., M.Luckett, M.Abdullah, J.C.H., Z.Z., H.Y., M.B., K.Haldar, E.T., M.Antoniou, F.M, M.J., C.Brookes, G.G., S.E., K.Hill, N.A.N., N.C.K., J.M., R.T., A.Kilgallon, A.Kulasinghe contributed to data analysis, interpretation, writing, and manuscript editing.

D.A.F., M.G.K., M.D., M.S., S.D., L.D., B.M., C.P., A.Bajaj, C.Belcher, D.F., J.L.L., M.Little, L.C. contributed to data collection.

## Supplementary Information

The online version contains supplementary material available at xxxxxx

## Supplementary Materials

Supplementary information Supplementary data tables MIST3 Protocol Source data

