## Supplementary Figures and Tables for "Tumour extrinsic neutrophil regulation in mesothelioma correlates with responsiveness to AXL and PD1 inhibition in MIST3, a phase IIA clinical trial"

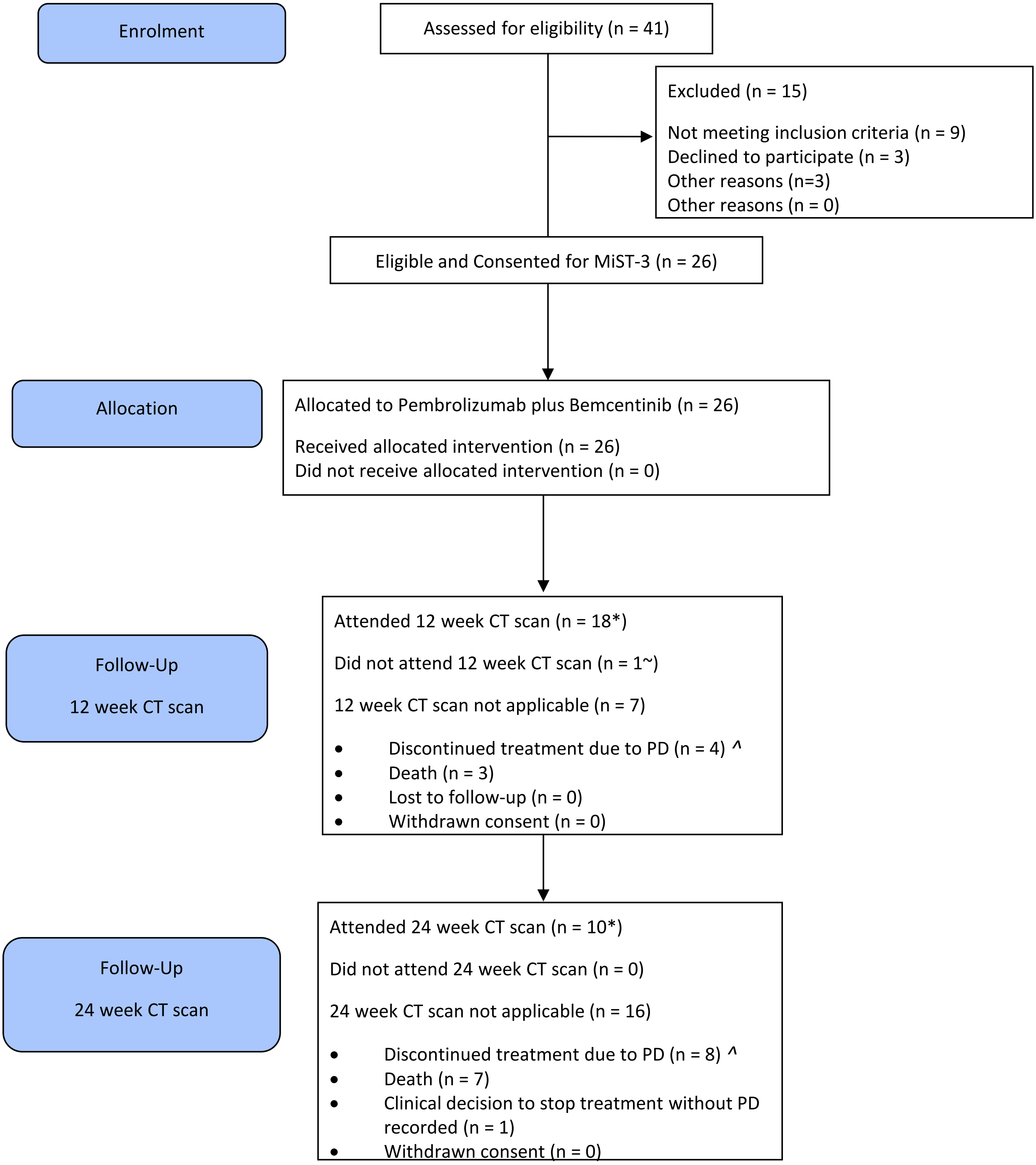


**Supplementary figure 1. CONSORT diagram for MiST3.**

*One of the 12 week CT scans was carried out later than the time window (13.1 weeks). Likewise one of the 24 week CT scans was carried out later than the time window (27.1 weeks). ~ This patient had a CT scan at 6 and 15.7 weeks and therefore has a 12 week CT scan response imputed for the primary analysis. ^includes clinical progressive disease (PD)

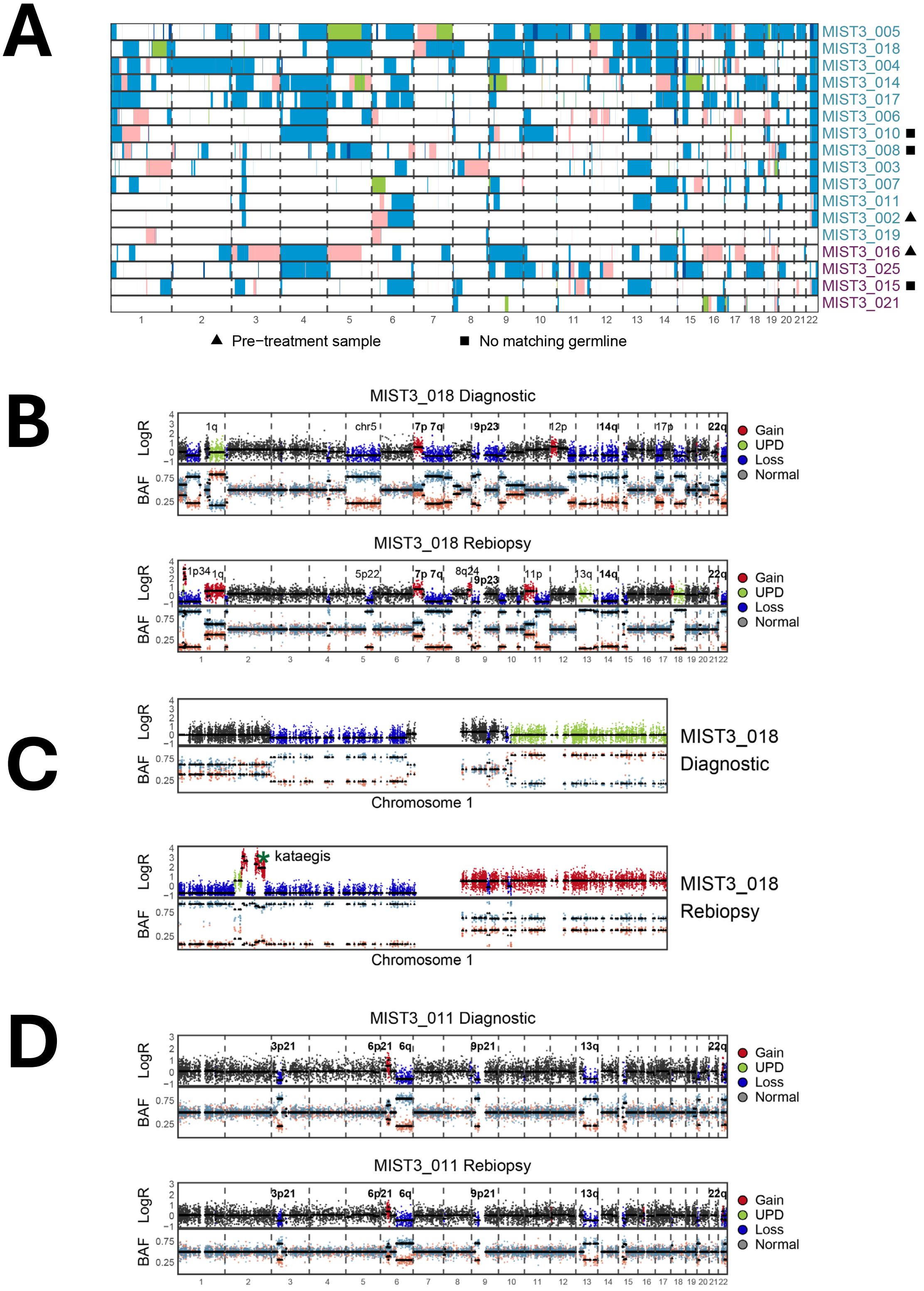


**Supplementary figure 2.**

A. Heat map showing the relative distribution of SCNAs in R (teal) versus NR (purple) patients.

B. Genome wide plots of BAF and log2 ratio in WES diagnostic and rebiopsy sample from patient MIST3_018, showing copy-number evolution between timepoints.

C. High resolution chromosome 1 plot of BAF and log2 ratio showing acquisition of kataegis with amplification of 1p34 at the time of disease progression in MIST3_018.

D. Genome wide plots of BAF and log2 ratio in WES diagnostic and rebiopsy samples from patient MIST3_011, showing stable losses of 9p21, 13q and 22q.

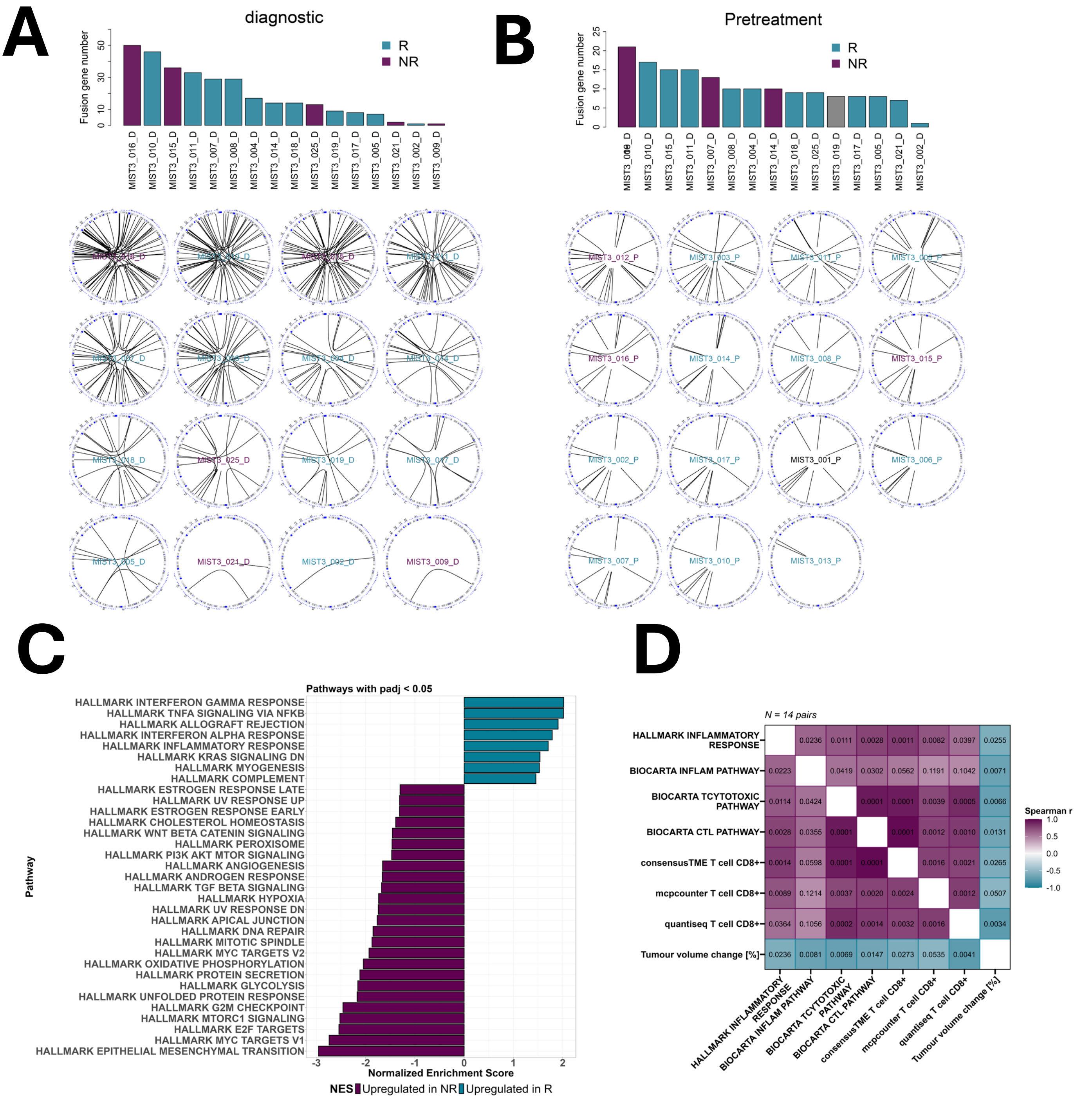


**Supplementary figure 3.**

A. Histogram showing the relative frequency of inferred translocations from bulk RNAseq analysis in diagnostic tissue. Corresponding circus plots related to translocations below.

B. Histogram showing the relative frequency of inferred translocations from bulk RNAseq analysis in pretreatment tissue. Corresponding circus plots related to translocations below.

C. Gene set enrichment analysis (GSEA) of the external ICB cohort (MIST4 + CONFIRM, N = 42) comparing R and NR using batch-adjusted diagnostic RNA-seq data. Bars show normalized enrichment scores (NES) for MSigDB HALLMARK pathways with BH-adjusted p < 0.05.

D. Spearman correlation matrix of best change in tumour size with CD8-related features and selected immune ssGSEA pathways from MIST3 pretreatment biopsies (N = 14). Tiles show correlation coefficients and corresponding p-values.

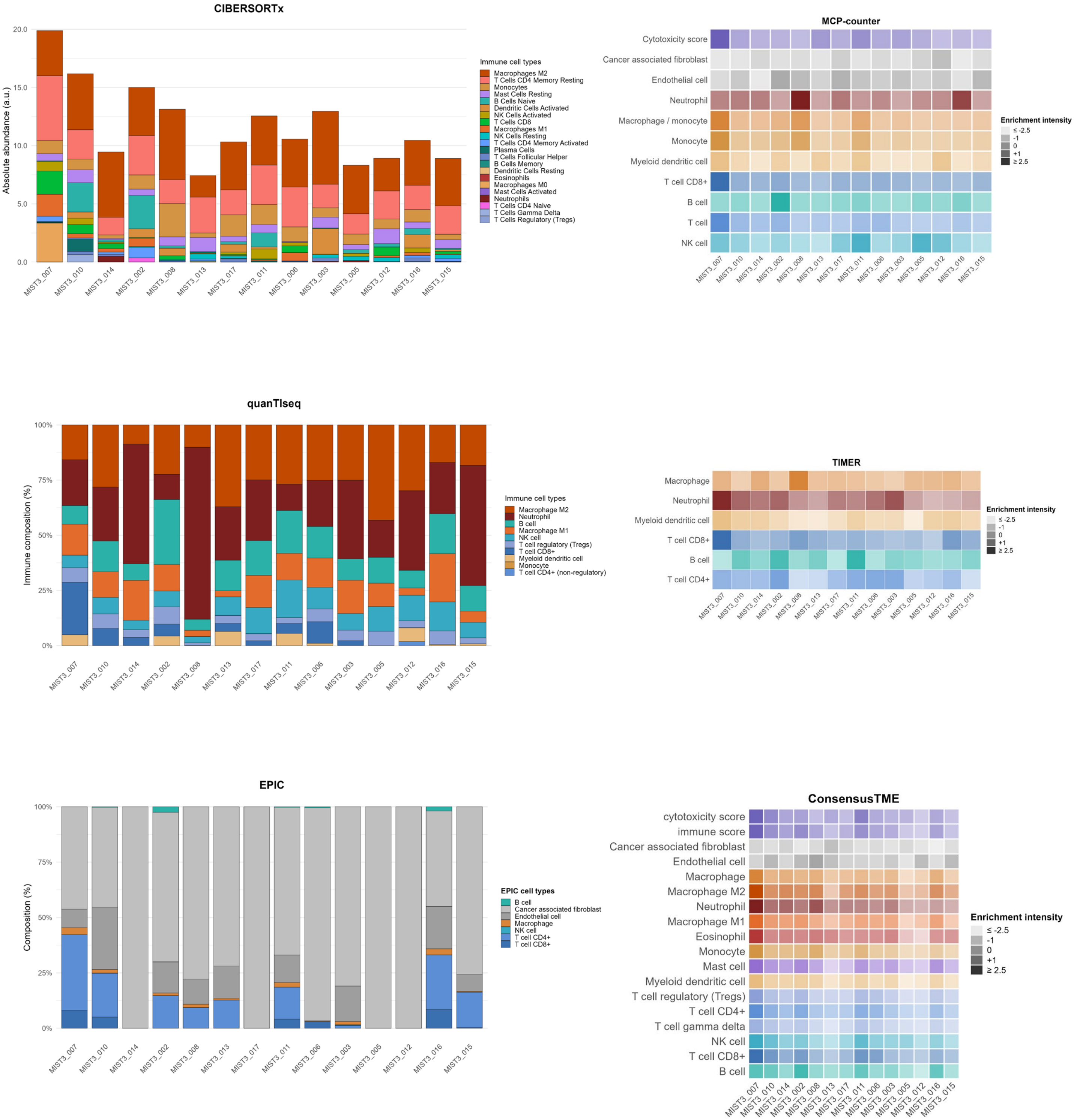


**Supplementary figure 4.**

Immune cell populations estimated by several deconvolution methods, visualized using method-appropriate scales (fractions vs. enrichment scores). Patients are ordered from left to right by best change in tumour size.

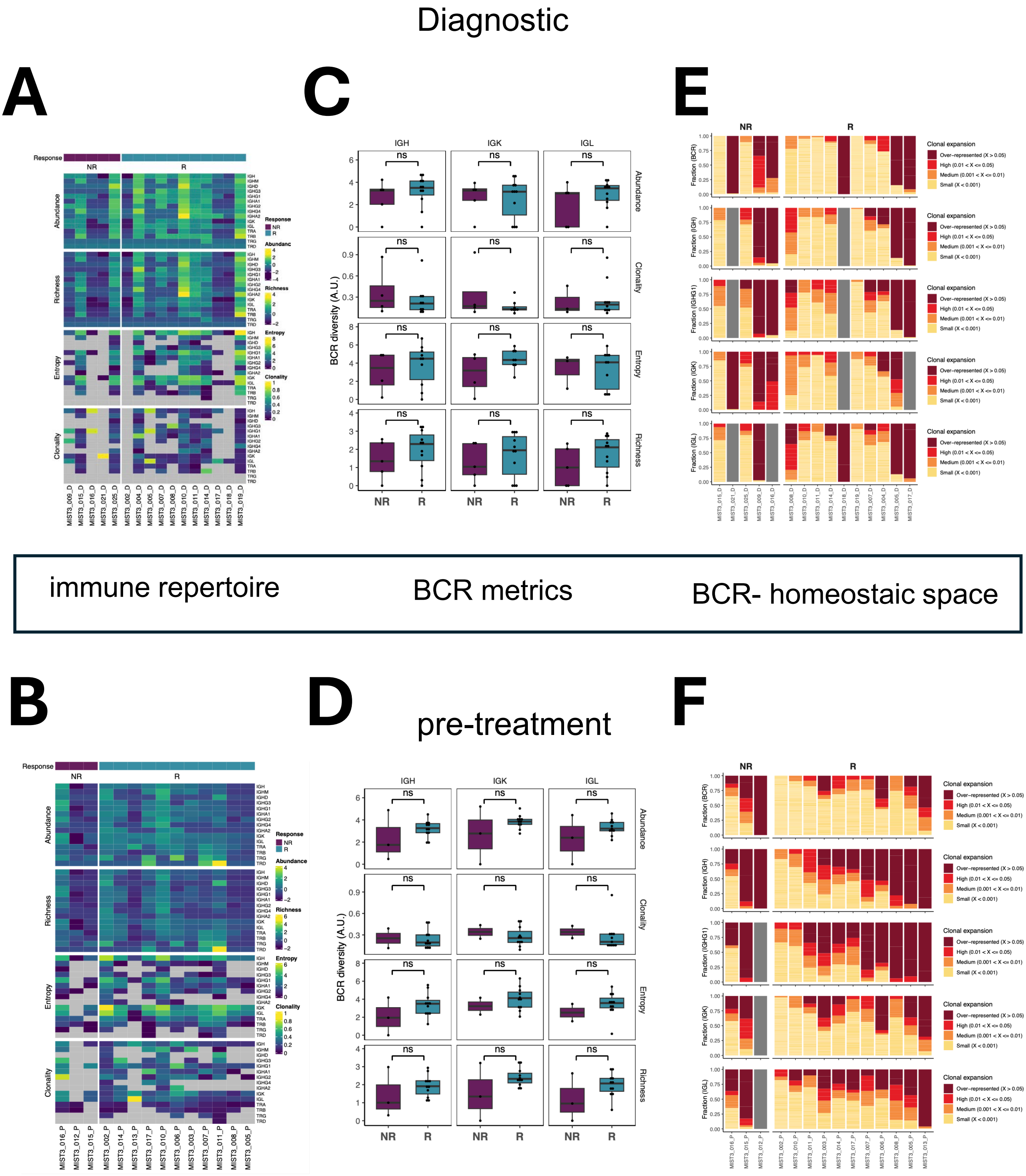


**Supplementary figure 5. Immune repertoire in the MIST3 clinical trial**
T cell receptor repertoire inferred from bulk RNA sequencing. A. Immune repertoire in diagnostic mesothelioma tissue. B. pretreatment tissue. Metrics corresponding to abundance, richness, entropy and clonality in the diagnostic C. and pre-treatement D. tissue. The R subgroup is shown as teal, and NR as purple. BCR metrics comparing R and NR subgroups in diagnostic E. and pre-treatment F. tissues (with respect to abundance, clonality, entropy, richness corresponding to immunoglobulin loci IGH, IGK and IGL)

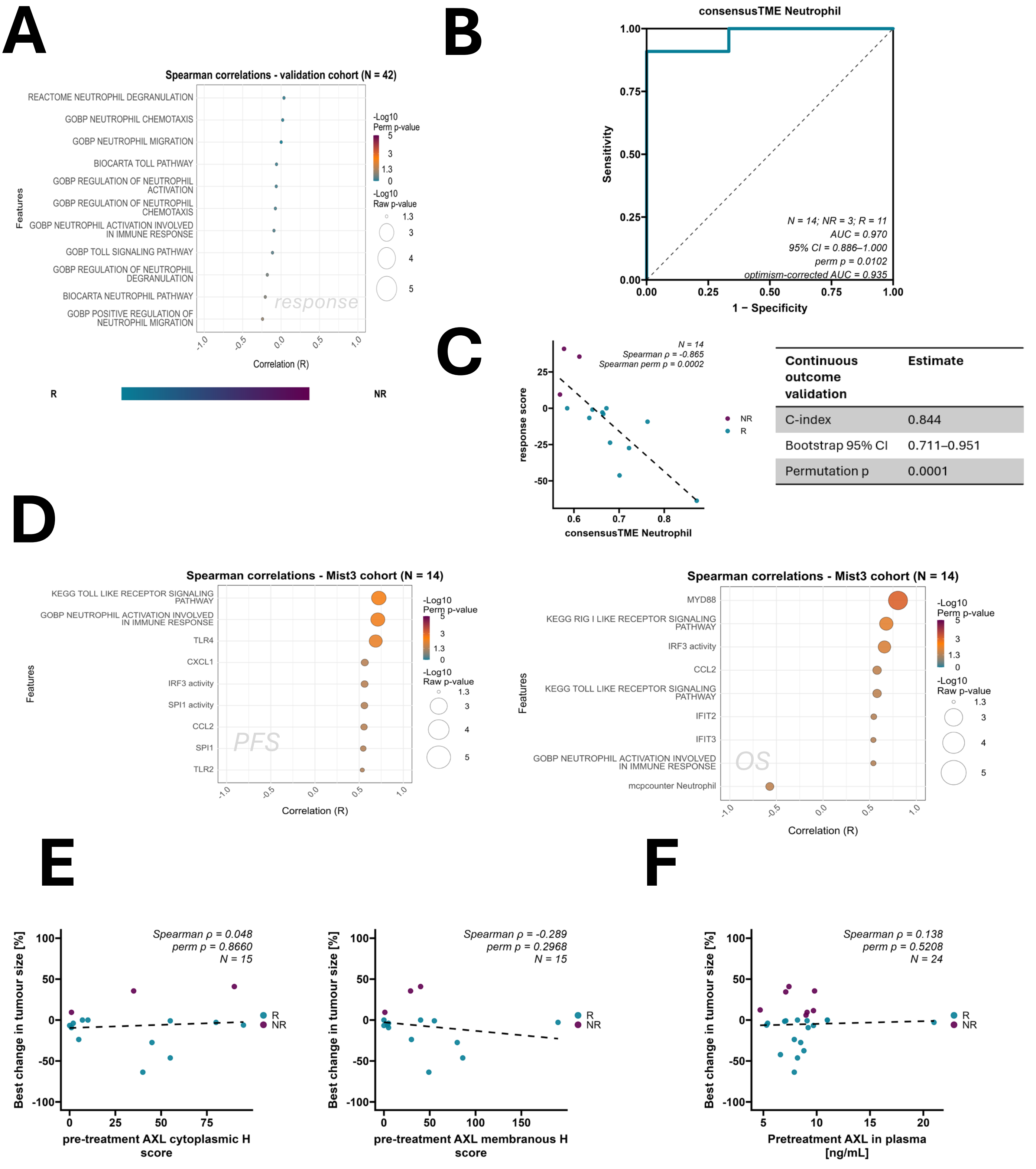


| Continuous outcome validation | Estimate |
|---|---|
| C-index | 0.844 |
| Bootstrap 95% CI | 0.711–0.951 |
| Permutation p | 0.0001 |

**Supplementary figure 6.**

A. Bubbleplot illustrating Spearman correlations of best change in tumour size with ssGSEA scores for neutrophil related processes in MiST4/Confirm (n = 42).
B. Exploratory ROC analysis of consensusTME neutrophil estimates for distinguishing R from NR. AUC, 95% CI, permutation p-value and optimism-corrected AUC are shown.
C. Association between consensusTME neutrophil score and continuous treatment response. Spearman correlation, permutation p-value and C-index are shown.
D. Bubbleplots showing Spearman correlations of chosen innate immunity related transcriptomic features with progression free and overall survival in pretreatment MIST3 biopsies.
E. Scatterplots showing correlations of best change in tumour size and AXL IHC results.
F. Scatterplot showing correlation of best change in tumour size and plasma level of AXL.

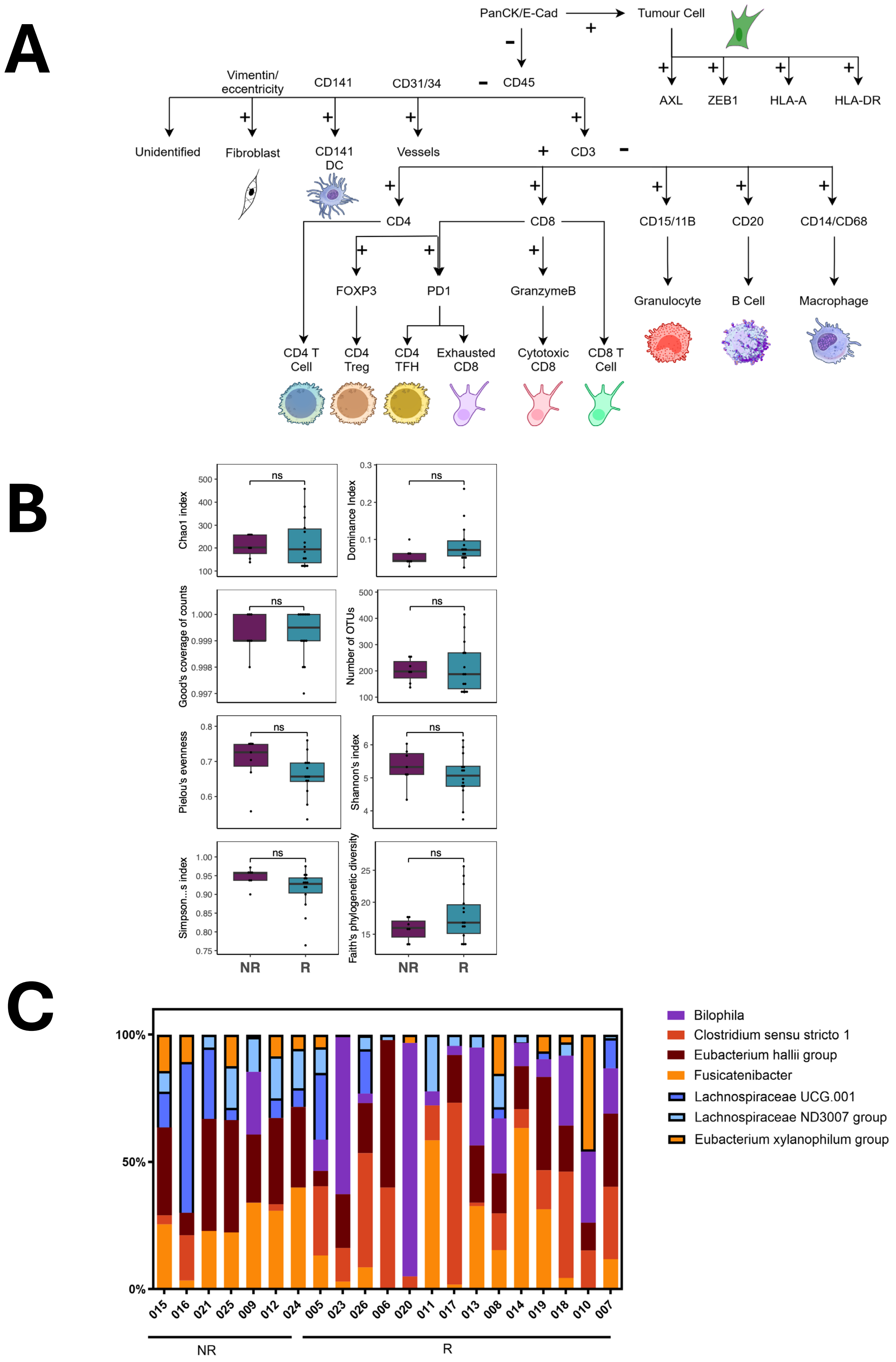


**Supplementary figure 7. Gut microbiota in the MIST3 clinical trial**

A. Schematic of cell phenotyping in 49-plex mfIHC staining.

B. Diversity metrics for R vs NR patients in the MIST3 cohort (NS = not significant by Mann Whitney U test, $p > 0.05$

C. Stacked histograms showing the relative, per patient abundance of significantly enriched genera in the MIST3 cohort

| Characteristics | Descriptive | Allocated to Pembrolizumab & Bemcentinib (n = 26) |
|---|---|---|
| Age (years) | Mean[SD] | 71.7 [5.8] |
| | Median[IQR] | 72.5 [69.0, 75.0] |
| | Min, max | 55.0, 85.0 |
| BMI (Kg/M$^2$) | Mean[SD] | 26.2 [4.1] |
| | Median[IQR] | 26.6 [23.4, 28.9] |
| | Min, max | 18.2, 32.7 |
| Gender | Male | 23 (88.5%) |
| | Female | 3 (11.5%) |
| Smoking status | Smoker | 1 (3.8%) |
| | Non-Smoker | 12 (46.2%) |
| | Ex-smoker | 13 (50.0%) |
| Mesothelioma subtype | Epithelioid | 23 (88.5%) |
| | Biphasic | 2 (7.7%) |
| | Sarcomatoid | 1 (3.8%) |
| History of asbestos | Yes | 20 (76.9%) |
| | Unknown | 6 (23.1%) |
| ECOG status | 0 | 6 (23.1%) |
| | 1 | 20 (76.9%) |
| Primary tumour site | Thoracic | 25 (96.2%) |
| | Missing | 1 (3.8%) |
| T-stage | Unobtainable | 2 (7.7%) |
| | T1 | 5 (19.2%) |
| | T2 | 2 (7.7%) |
| | T3 | 9 (34.6%) |
| | T4 | 6 (23.1%) |
| | TX | 2 (7.7%) |
| N-stage | Unobtainable | 2 (7.7%) |
| | N0 | 16 (61.5%) |
| | N1 | 6 (23.1%) |
| | N2 | 2 (7.7%) |
| M-stage | Unobtainable | 2 (7.7%) |
| | M0 | 19 (73.1%) |
| | M1 | 5 (19.2%) |

**Supplementary table 1** showing baseline patient characteristics. Data are n (%), median [IQR], Mean [SD], or min and max; Abbreviations: BMI=Body Mass Index; ECOG=Eastern Cooperative Oncology Group; NOS=Not Otherwise Specified; IQR=Inter-quartile range; SD=Standard deviation

| Characteristics | Descriptive | Total allocated (n = 26) |
|---|---|---|
| Number of prior courses of systemic anticancer therapy | One | 17 (65.4%) |
| | Two | 7 (26.9%) |
| | Three | 2 (7.7%) |
| Best response of first line therapy | Partial response | 1 (3.8%) |
| | Stable disease | 13 (50.0%) |
| | Progressive disease | 12 (46.2%) |
| First line therapy | Pemetrexed and/or Carboplatin | 11 (42.3%) |
| | Pemetrexed and/or Cisplatin | 8 (30.8%) |
| | Radiotherapy | 1 (3.8%) |
| | Pemetrexed and Cisplatin/Carboplatin | 1 (3.8%) |
| | Bevacizumab and Pemetrexed and/or Carboplatin | 5 (19.2%) |
| Second line therapy | Pemetrexed and/or Carboplatin | 3 (33.3%) |
| | Gemcitabine & AZD6738 | 1 (11.1%) |
| | Pemetrexed/Carboplatin and Bevicizumab | 1 (11.1%) |
| | Bevacizumab | 2 (22.2%) |
| | Vinorelbine & carboplatin | 1 (11.1%) |
| | Radiotherapy | 1 (11.1%) |
| Third line therapy | Vinorelbine | 1 (50%) |
| | Carboplatin and Pemetrexed | 1 (50%) |

**Supplementary table 2**
Summary of patients’ previous courses of systemic anticancer therapy

| Participant ID | Time on treatment (weeks) | Reason for discontinuation | Provided 12 week CT scan | Provided 24 week CT scan |
|---|---|---|---|---|
| **MIST3_001** | 1.7 | Clinical decision | No | No |
| **MIST3_017** | 4.6 | Disease Progression£ | No | No |
| **MIST3_015** | 6.6 | Disease Progression£ | No | No |
| **MIST3_003** | 7.3 | Disease Progression£ | No | No |
| **MIST3_022** | 8.3 | Clinical decision | No | No |
| **MIST3_021** | 8.3 | Disease Progression£ | No | No |
| **MIST3_020** | 9.3 | Clinical decision | No | No |
| **MIST3_026** | 10.1 | Clinical decision | Yes[a] | No |
| **MIST3_023** | 13 | Disease Progression£ | Yes | No |
| **MIST3_025** | 13.3 | Disease Progression£ | Yes | No |
| **MIST3_014** | 13.3 | Disease Progression£ | Yes | No |
| **MIST3_005** | 13.6 | Disease Progression£ | Yes | No |
| **MIST3_009** | 14 | Disease Progression£ | Yes | No |
| **MIST3_016** | 14.1 | Disease Progression£ | Yes | No |
| **MIST3_012** | 14.3 | Disease Progression£ | Yes | No |
| **MIST3_006** | 19.3 | Disease Progression£ | Yes | No |
| **MIST3_008** | 27.7 | Clinical decision | Yes | Yes |
| **MIST3_010** | 37.1 | Disease Progression^ | Yes | Yes |
| **MIST3_013** | 37.3 | Disease Progression^ | Yes | Yes |
| **MIST3_011** | 37.3 | Disease Progression^ | Yes | Yes |
| **MIST3_018** | 38.3 | Disease Progression^ | Yes | Yes |
| **MIST3_024** | 40.3 | Disease Progression^ | Yes | Yes |
| **MIST3_004** | 50.4 | Disease Progression^ | Yes | Yes |
| **MIST3_019** | 53.3 | Clinical decision | Yes | Yes |
| **MIST3_002** | 79.4 | Disease Progression£ | Yes | Yes |
| **MIST3_007** | 106.4[b] | N/A[b] | Yes | Yes |

[a] 12 week CT scan not taken but imputed from CT scan before and after
[b] This patient remained on treatment at data lock, time on treatment relates to their cycle 35 treatment date which was last cycle received prior to data lock.
^Clinical disease progression
£CT scan confirmed disease progression

**Supplementary Table 3**
Summary of patients' times on study with the reason for discontinuation

**A**

| | | MiST-3 (n = 26)<br><br>n (%) |
|---|---|---|
| **Patients with any AEs** | | 26 (100%) |
| | Patients with one AE | 2 (7.7%) |
| | Patients with two AEs | 2 (7.7%) |
| | Patients with three AEs | 3 (11.5%) |
| | Patients with four AEs | 1 (3.8%) |
| | Patients with five or more AEs | 18 (69.2%) |
| **Patients without AEs** | | 0 (0%) |
| **Patients with grade 3 and above AEs** | | 14 (53.9%) |

**B**

| | | MiST-3 (n = 26)<br><br>n (%) |
|---|---|---|
| Total AEs | | 248 |
| By CTCAE grade | | |
| | Missing | 2 (0.8%) |
| | 1 | 171 (69.0%) |
| | 2 | 57 (23.0%). |
| | 3 | 18 (7.3%) |
| | 4 | 0 (0.0%) |
| | 5 | 0 (0.0%) |
| By relatedness to Pembrolizumab | | |
| | Missing | 2 (0.8%) |
| | Not Related | 117 (47.2%) |
| | Unlikely | 45 (18.2%) |
| | Possibly | 51 (20.6%) |
| | Probably | 14 (5.7%) |
| | Definitely | 19 (7.7%) |
| By relatedness to Bemcentinib | | |
| | Missing | 2 (0.8%) |
| | Not Related | 121 (48.8%) |
| | Unlikely | 24 (9.7%) |
| | Possibly | 68 (27.4%) |
| | Probably | 13 (5.2%) |
| | Definitely | 20 (8.1%) |

**Supplementary Table 4 Adverse Events (AEs)**

A. Number of AEs per patient. Data are presented as number (n) and percentage (%) of patients
B. AEs by CTCAE grade and relatedness to pembrolizumab and bemcentinib. Data are presented as number (n) and percentage (%) of AEs, calculated using the number of AEs in each row divided by the total number of AEs reported.

| | | MiST-3 (n = 26)<br>n (%) |
|---|---|---|
| **Number of recruited participants in MiST-3** | | 26 (100%) |
| **By CTCAE grade** | | |
| | Grade 1 | 5 (19.2%) |
| | Grade 2 | 7 (26.9%) |
| | Grade 3 | 14 (53.8%) |
| | Grade 4 | 0 (0.0%) |
| | Grade 5 | 0 (0.0%) |
| **By relatedness to Pembrolizumab** | | |
| | Not Related | 4 (15.4%) |
| | Unlikely | 2 (7.7%) |
| | Possibly | 8 (30.8%) |
| | Probably | 4 (15.4%) |
| | Definitely | 8 (30.8%) |
| **By relatedness to Bemcentinib** | | |
| | Not Related | 6 (23.1%) |
| | Unlikely | 1 (3.8%) |
| | Possibly | 6 (23.1%) |
| | Probably | 6 (23.1%) |
| | Definitely | 7 (26.9%) |
| **By CTCAE grade for TRAE to Pembrolizumab** | | |
| | Not TRAE | 6 (23.1%) |
| | TRAE Grade 1 | 12 (46.2%) |
| | TRAE Grade 2 | 2 (7.7%) |
| | TRAE Grade 3 | 6 (23.1%) |
| | TRAE Grade 4 | 0 (0.0%) |
| | TRAE Grade 5 | 0 (0.0%) |
| **By CTCAE grade for TRAE to Bemcentinib** | | |
| | Not TRAE | 7 (26.9%) |
| | TRAE Grade 1 | 7 (26.9%) |
| | TRAE Grade 2 | 6 (23.1%) |
| | TRAE Grade 3 | 6 (23.1%) |
| | TRAE Grade 4 | 0 (0.0%) |
| | TRAE Grade 5 | 0 (0.0%) |

**Supplementary Table 5 Adverse Events (AEs) by grade and relatedness**. Highest level of AEs by grade and relatedness experience by each participant, where TRAEs are defined as those possibly, probably or definitely related. Data are presented as number (n) and percentage (%) of patients. TRAE, treatment-related adverse events.

| | MiST-3 (n = 26) | | | | | |
|---|---|---|---|---|---|---|
| | Any grade | Split by individuals' highest CTCAE grade | | | | |
| | | Grade 1 | Grade 2 | Grade 3 | Grade 4 | Grade 5 |
| **Any AEs** | 26 (100%) | 5 (19%) | 7 (27%) | 14 (53.9%) | 0 (0%) | 0 (0%) |
| **AEs by preferred term (for AEs experienced by ≥10% of patients or with at least one event of CTCAE≥3)** | | | | | | |
| Fatigue | 12 (46%) | 10 (38%) | 1 (4%) | 1 (4%) | 0 (0%) | 0 (0%) |
| Nausea | 11 (42%) | 11 (42%) | 0 (0%) | 0 (0%) | 0 (0%) | 0 (0%) |
| Constipation[a] | 7 (27%) | 6 (23%) | 0 (0%) | 0 (0%) | 0 (0%) | 0 (0%) |
| Weight loss | 7 (27%) | 3 (12%) | 3 (12%) | 1 (4%) | 0 (0%) | 0 (0%) |
| Diarrhoea | 7 (27%) | 6 (23%) | 0 (0%) | 1 (4%) | 0 (0%) | 0 (0%) |
| Raised creatinine | 6 (23%) | 6 (23%) | 0 (0%) | 0 (0%) | 0 (0%) | 0 (0%) |
| Swollen legs and feet | 5 (19%) | 5 (19%) | 0 (0%) | 0 (0%) | 0 (0%) | 0 (0%) |
| Increased ALT | 5 (19%) | 4 (15%) | 1 (4%) | 0 (0%) | 0 (0%) | 0 (0%) |
| Increased AST | 5 (19%) | 5 (19%) | 0 (0%) | 0 (0%) | 0 (0%) | 0 (0%) |
| Anaemia | 5 (19%) | 1 (4%) | 4 (15%) | 0 (0%) | 0 (0%) | 0 (0%) |
| Back Pain | 4 (15%) | 3 (12%) | 1 (4%) | 0 (0%) | 0 (0%) | 0 (0%) |
| Dyspnoea | 4 (15%) | 1 (4%) | 3 (12%) | 0 (0%) | 0 (0%) | 0 (0%) |
| Rash | 4 (15%) | 3 (12%) | 0 (0%) | 1 (4%) | 0 (0%) | 0 (0%) |
| Fever | 4 (15%) | 2 (8%) | 2 (8%) | 0 (0%) | 0 (0%) | 0 (0%) |
| Decreased appetite | 3 (12%) | 2 (8%) | 1 (4%) | 0 (0%) | 0 (0%) | 0 (0%) |
| Joint pain | 3 (12%) | 2 (8%) | 0 (0%) | 1 (4%) | 0 (0%) | 0 (0%) |
| Acid reflux | 3 (12%) | 2 (8%) | 1 (4%) | 0 (0%) | 0 (0%) | 0 (0%) |
| Hypomagnesemia | 3 (12%) | 3 (12%) | 0 (0%) | 0 (0%) | 0 (0%) | 0 (0%) |
| Oral Candidiasis | 3 (12%) | 0 (0%) | 2 (8%) | 1 (4%) | 0 (0%) | 0 (0%) |
| shortness of breath | 3 (12%) | 1 (4%) | 2 (8%) | 0 (0%) | 0 (0%) | 0 (0%) |
| Urinary tract infection | 2 (8%) | 1 (4%) | 0 (0%) | 1 (4%) | 0 (0%) | 0 (0%) |
| Hypophosphatemia | 2 (8%) | 1 (4%) | 0 (0%) | 1 (4%) | 0 (0%) | 0 (0%) |
| Arthralgia | 1 (4%) | 0 (0%) | 0 (0%) | 1 (4%) | 0 (0%) | 0 (0%) |
| Hyponatremia | 1 (4%) | 0 (0%) | 0 (0%) | 1 (4%) | 0 (0%) | 0 (0%) |
| Pneumonitis | 1 (4%) | 0 (0%) | 0 (0%) | 1 (4%) | 0 (0%) | 0 (0%) |
| Community acquired pneumonia | 1 (4%) | 0 (0%) | 0 (0%) | 1 (4%) | 0 (0%) | 0 (0%) |
| Empyema | 1 (4%) | 0 (0%) | 0 (0%) | 1 (4%) | 0 (0%) | 0 (0%) |
| Thrombocytopenia | 1 (4%) | 0 (0%) | 0 (0%) | 1 (4%) | 0 (0%) | 0 (0%) |
| Hypercalcaemia | 1 (4%) | 0 (0%) | 0 (0%) | 1 (4%) | 0 (0%) | 0 (0%) |
| Autoimmune hepatitis | 1 (4%) | 0 (0%) | 0 (0%) | 1 (4%) | 0 (0%) | 0 (0%) |
| Gallstones | 1 (4%) | 0 (0%) | 0 (0%) | 1 (4%) | 0 (0%) | 0 (0%) |
| Colitis | 1 (4%) | 0 (0%) | 0 (0%) | 1 (4%) | 0 (0%) | 0 (0%) |

**Supplementary table 6.** Treatment emergent adverse events that either happened in at least 3 patients (>= 10%) or were grade 3 or above by CTCAE grade and preferred term

[a] Highest grade columns does not add to total having any grade as one individual has grade missing.

# A

| | MiST-3 (n = 26)<br><br>n (%) |
|---|---|
| **Patients with SAEs** | **10 (38.5%)** |
| Patients with one SAE | 9 (34.6%) |
| Patients with two SAEs | 1 (3.9%) |
| Patients with three SAEs | 0 (0%) |
| Patients with more than three SAEs | 0 (0%) |
| **Patients without SAEs** | **16 (61.5%)** |
| Patients with SAE leading to treatment discontinuation^ | 10 (38.5%) |
| Patients with SAE leading to trial withdrawal | 0 (0%) |
| Patients with SAE leading to patient death | 0 (0%) |

# B

| **Any SAEs** | **n = 11** |
|---|---|
| Related to Pembrolizumab | |
| Yes | 3 (27.3%) |
| No | 8 (72.7%) |
| Related to Bemcentinib | |
| Yes | 3 (27.3%) |
| No | 8 (72.7%) |
| Final grade of SAE | |
| Grade 1 | 1 (9.1%) |
| Grade 2 | 3 (27.3%) |
| Grade 3 | 7 (63.7%) |
| Grade 4-5 | 0 (0%) |
| Final outcome | |
| Resolved | 10 (90.9%) |
| Missing | 1 (9.1%) |
| Fatal | 0 (0%) |

**Supplementary Table 7**

A. Number of serious adverse events (SAEs) per patient. ^if at least one of the pembrolizumab or bemcentinib was temporarily discontinued, then it is considered as treatment discontinuation. Data are presented as number (n) and percentage (%) of participants, SAE; Serious adverse events

B. Serious adverse events (SAEs) by CTCAE grade and relatedness to pembrolizumab and bemcentinib. Data are presented as number (n) and percentage (%) of SAEs

| Trial ID | SAE term | CTCAE Grade | Bemcentinib Causality | Pembrolizumab Causality | Outcome |
|---|---|---|---|---|---|
| MIST3_008 | Fever | 1 | Definitely | Definitely | Resolved |
| MIST3_010 | Fever | 2 | Unrelated | Unrelated | Resolved |
| MIST3_004 | Thrombocytopenia | 3 | Definitely | Unlikely | Missing |
| MIST3_018 | Empyema | 3 | Unrelated | Unrelated | Resolved |
| MIST3_022 | Community Acquired Pneumonia | 3 | Unrelated | Unrelated | Resolved |
| MIST3_021 | Urinary Tract Infection | 3 | Unrelated | Unrelated | Resolved |
| MIST3_023 | Dyspnoea | 2 | Unrelated | Unrelated | Resolved |
| MIST3_025 | Sepsis | 2 | Unrelated | Unrelated | Resolved |
| MIST3_023 | Pneumonitis | 2 | Unrelated | Definitely | Resolved |
| MIST3_026 | Autoimmune Hepatitis | 3 | Definitely | Definitely | Resolved |
| MIST3_019 | Gallstones | 3 | Unrelated | Unrelated | Resolved |

**Supplementary Table 8**
Description of serious advent events (SAEs) per patient.
